# Mixed methods evaluation of the “train the trainer” model for delivering core “Making Every Contact Count” (MECC) training

**DOI:** 10.64898/2025.12.11.25342064

**Authors:** Beth Nichol, Angela M. Rodrigues, Mei Yee Tang, Anna Haste, Sarah Audsley, Craig Robson, Jill Harland, Catherine Haighton

## Abstract

**Introduction:** The current study aimed to evaluate the regional Implementation of the Train the Trainer (TtT) model to deliver Making Every Contact Count (MECC, an opportunistic and person-centered approach to the promotion of health and wellbeing) training across the North East and North Cumbria.

**Methods:** A mixed methods evaluation included secondary data from pre and post MECC TtT training survey evaluations and cascade behaviour of MECC trainers, qualitative semi-structured interviews with individuals who were eligible for the MECC TtT programme (n = 21) analysed according to the Theoretical Domains Framework (TDF), content analysis of the existing MECC TtT programme to identify behaviour change techniques (BCTs) and intervention functions (IFs), and a strategic behavioural analysis to identify the extent to which the current programme addressed the behavioural problem.

**Results:** Only 4.4% of MECC trainers who completed the post survey evaluation reported cascade of core MECC training following the TtT programme. MECC TtT training significantly improved knowledge but not confidence or motivation of trainees to deliver the MECC training. However, motivation and confidence post training significantly predicted intention to cascade. Neither mode of delivery or post training knowledge significantly predicted intention to cascade. Six key TDF domains were identified as key barriers and facilitators to cascade of core MECC training; Environmental Context and Resources, Knowledge, Social Influences, Beliefs about Consequences, Skills, and Intentions. MECC TtT training only sufficiently addressed the key TDF domain Skills.

**Conclusions:** A targeted approach to recruitment to the regional MECC TtT programme should be adopted to ensure trainees have top-down and peer support, previous MECC knowledge and delivery within their role and experience of delivering training. Otherwise, additional support is needed for MECC trainers.

## Background

Making Every Contact Count (MECC) is defined by its person-centred approach to promoting health and wellbeing during everyday conversations [1], enabling individuals to engage in conversations about their health across multiple organisations and populations [2]. The topic and duration of a MECC conversation and the specific skills and techniques applied are determined by the needs of the individual [1]. For example, this may include conversations around smoking, alcohol, physical activity, or diet to motivate individuals to make a behaviour change [1], or assist individuals around the wider determinants such as debt management, housing, and employment by providing advice and signposting to services for further support [3]. Given that MECC does not require specialist skills, anyone could potentially deliver MECC [1]. Thus, MECC poses relevance to organisations across the public, private, and third sectors. Although a paucity of literature has investigated the effectiveness of MECC in improving health and wellbeing of service users, there is some evidence to indicate that MECC may increase dietary quality and decrease sedentary time [4]. Furthermore, the rationale for MECC draws upon the evidence base to support comparable interventions such as brief motivational interviewing [5, 6], which employs similar principles of motivating individuals to make changes to their behaviour [7]. More frequently, MECC literature reports that receipt of MECC training significantly increases the confidence [8–11] and competence [10–15] of trainees to deliver MECC to service users, and that MECC is generally acceptable to service providers [8, 13, 16] and users [14, 17] who report that they feel listened to [12, 17] and thus are more satisfied with interactions compared to usual care [12].

To facilitate the delivery of MECC at scale, a Train the Trainer (TtT) model of MECC training is often adopted for regional implementation of MECC [18]. The TtT model aims to equip recipients of TtT training with the knowledge and skills to become trainers, cascading training to frontline staff thus creating a ‘ripple effect’ [19].

Theoretically when compared to the traditional model of training delivery by professional trainers, the TtT model facilitates training delivery across sectors with the greatest cost efficiency and with the potential to reach a larger number of individuals [20, 21]. Additionally, assuming recipients of TtT are local to the audiences they will train, the TtT model facilitates training delivery that is sensitive to the audience and the community it is delivered within [20, 21]. Building capacity at the local level also has the potential for enhancing collaboration and networking among those trained and for sustaining the training [22]. Thus, the TtT model provides a viable approach to dissemination and scale up of public health practices [20]. When defining effectiveness of the TtT model, it is important to consider both the ability of the model to deliver training (and ultimately the target behaviour(s) taught during training) to service users at scale, but also the quality of the training that is provided (and subsequently the quality of the intervention that is delivered).

Studies of successful implementation of the TtT model indicate both; training delivery can reach a larger number of individuals if cascaded successfully [21], and the vast majority of recipients of training delivered by TtT trainers report high levels of quality of and satisfaction with the training [23, 24], and that training helped them to implement the target behaviour(s) [20, 25]. Furthermore, another study found utility of training in implementing the target behaviour(s) to be comparable to when delivered by experts, showing the same increases in knowledge, intention to perform the behaviour, ability, and motivation [26]. A study exploring the TtT model for MECC training delivery also identified that training significantly reduced barriers to cascade [27].

The success of implementation of the TtT model varies considerably, with the proportion of TtT trainees reported to have cascaded training ranging from 20% [22, 25] to 96% [21]. Reported barriers to cascading training include resource restrictions, staff movement and organisational change, and insufficient confidence [24]. The available MECC literature indicates similar challenges in implementing the TtT model due to barriers including limited staff capacity [18] and motivation [28] to deliver and attend training. Facilitators to the implementation of a TtT model of MECC training include adequate training resources, motivated trainees, top-down support, sustained funding, and a culture change within organisations towards promoting MECC [28], although a formal evaluation of the implementation of the TtT model specifically is lacking.

Despite its widespread use and potential benefits, the literature on the effectiveness of TtT models is limited, as existing studies often only measure intentions to deliver training after TtT training [29] rather than evidence of training cascade. Particularly, there is also a paucity of literature examining the TtT model in the context of MECC, despite its current widespread implementation. To evaluate the current TtT model to deliver MECC training, a holistic approach using mixed methods allowed for triangulation of qualitative and quantitative data [30], in keeping with the Medical Research Council (MRC) Framework for evaluation of complex interventions [31].

Quantitative data can inform fidelity to the intervention, which refers to the delivery of an intervention as intended [32], in this case the receipt of the intervention (changes in ability pre and post MECC TtT training) and the outcomes of the intervention (rates of cascade of MECC training). Concurrently, use of behaviour change theory can provide a framework to evaluate training, as training that is based within health psychology improves recipients’ confidence, competence, and intention to conduct health behaviour change conversations such as MECC [33].

Specifically, the behaviour change wheel can be applied to evaluate and optimise existing interventions [34]. The Theoretical Domains Framework (TDF) describes 14 domains of behavioural determinants that are congruent with capability, opportunity, or motivation to perform a behaviour and provides utility in understanding barriers and facilitators to performing the target behaviour [35]. Intervention functions, which describe broad categories of strategies of interventions [34], and BCTs, which specify 93 active components of an intervention [36], can be used to evaluate existing interventions. Both can be mapped onto the TDF to assess the extent to which the current intervention sufficiently addressed barriers to the intervention or whether there are missed opportunities for optimisation, through a strategic behavioural analysis (SBA) method [37]. Although a previous quantitative study utilised the TDF to detect how MECC TtT training using the Healthy Conversation Skills (HCS) addressed barriers and facilitators to training cascade [27], HCS is a specific approach to MECC TtT training which is non-didactic and includes practice and emphasises reflection [27]. By contrast, regional implementation within other areas including the North East and North Cumbria (NENC) does not possess the same theoretical underpinnings and thus less is known about implementation of MECC TtT that is not based within HCS. Therefore, this study aimed to use mixed methods to complete a comprehensive evaluation of the regional TtT model of delivering MECC training and provide recommendations to enhance uptake of the course and maximise delivery of core MECC to frontline staff across sectors.

## Methods

### Study design and setting

This evaluation adopted a concurrent mixed methods design [38], combining the following primary and secondary data collection methods:

1. Analysis of existing quantitative data collected before and after MECC TtT training (secondary data).
2. Analysis of qualitative data including in-depth qualitative interviews with MECC trainers and staff eligible for TtT in the NENC (primary data) and free text responses within the post MECC TtT survey using the TDF [35]. This stage is described in full within the qualitative manuscript [39].
3. Content analysis of the MECC TtT programme using the behaviour change technique taxonomy (v1) [36] (secondary data).
4. Triangulated in a Strategic Behavioural Analysis; namely assessment of the suitability of the MECC TtT programme and opportunities for improvement through mapping the BCTs it utilised onto the barriers and facilitators to cascading MECC training identified using the TDF.

The study protocol was preregistered prior to data collection on Open Science Framework (https://osf.io/xz8au). Reporting of this mixed methods study adhered to the Mixed Methods Article Reporting Standards (MMARS) outlined by the American Psychological Association (APA) Publications and Communications Board Working Group on Journal Article Reporting Standards (JARS) [40].

This study focused on the NENC region in England, where the TtT model of MECC training delivery has been formally adopted since 2022 to maximise delivery of core MECC at scale across sectors. Core MECC training is considered the foundational understanding of MECC and includes the rationale behind MECC, the concept of MECC, how to deliver MECC, and two to three example topics (e.g mental health, smoking, or alcohol); the specific selected topics tailored to the audience. Since 2022, the TtT model has been the primary strategy of implementation of core MECC training on a regional level, although core MECC training is still provided. Training is coordinated by the MECC regional at scale coordinator and delivered by the highly experienced MECC at Scale TtT course trainer (principal trainer). Training is advertised across MECC groups and other networks, and individuals from organisations across public, private, and third sectors are able to attend at no cost to the individual or their organisation. For example, recipient settings of MECC TtT training include local authority, NHS Trusts, charities, criminal justice settings, and Natural England. Over a 3.5-hour session, the MECC TtT course aims to enable trainees to become MECC trainers and deliver the core MECC training to their front-line colleagues in their organisations, or ‘cascade’ MECC training. MECC TtT training in the NENC adopts a passive and didactic pedagogy reminiscent of the ‘see one, do one, teach one’ concept within medical training [41]. Thus, the MECC course is taught as a demonstration of its delivery followed by specific advice and further information, with some opportunities for MECC trainers to customise slides, videos, and activities to the intended audience. The scope of this study covers all MECC trainers and staff eligible for TtT in the NENC, across all sectors MECC is offered to in this area.

This study was approved by the Ethics department at *(blinded for peer review)* This study also received R&D approval from the North East and North Cumbria Integrated Care System (ICS) as a service evaluation, as determined by the Health Research Authority.

### Professional contributor involvement

Four professional contributors, recruited via existing contacts and from MECC stakeholder organisations across NENC, represented local authority (n = 1), healthcare (n = 1), and the third sector (n = 2) formed an advisory panel. Any individuals eligible for MECC TtT training could be included on the professional contributor panel, whether they had received MECC TtT training or not. The panel met twice over the course of the research project, before and after data collection. In the first meeting, the panel advised on the study objectives, topic guides, and strategies for recruitment for participant interviews, and subsequently amendments were made to the topic guide. During the second meeting, panelists provided their feedback and interpretation on the study findings, informed on the lay summary, and advised on the feasibility of the recommendations made for MECC TtT training using the APEASE criteria, which outlines an acronym to assess whether a recommendation is Acceptable, Practical, Effective, Affordable, Safe and without unwanted side effects, and Equitable across all relevant stakeholders [34]. Consequently, the recommendations were amended to improve their applicability to practice.

#### 1. Secondary analysis of quantitative data

##### Source of data

Secondary data was received from the MECC regional at scale coordinator and included all data from the NENC network, recorded via the MECC Gateway app, relating to MECC TtT training from May 2022 to 2^nd^ November 2023 (point of retrieval). MECC TtT trainers were asked to complete a pre and post evaluation survey before and after attending the training, respectively. Anonymised participant IDs allowed pre and post-survey responses to be matched. Occasionally, more than one pre or post survey had been completed by the same participant. In this case, only the pre and post surveys with the same recorded date were retained. For individuals that responded to the pre and post survey, there was no missing data. MECC trainers were asked to conduct pre and post evaluation surveys when cascading MECC training. The responses of attendees that completed the surveys could be matched to the unique MECC trainer via an anonymous ID number. Evaluation surveys included the date thus both the number of training sessions and the number of individuals trained by each MECC trainer could be calculated. Therefore, data was both longitudinal (pre and post MECC TtT evaluation surveys) and cross-sectional (fidelity to cascade MECC training).

Participants were adults (≥ 18) currently residing in the UK that had received the MECC TtT programme who provided opt-in consent for their data to be used for evaluation purposes. The secondary data initially included 651 pre-survey and 373 post-survey responses. Once duplicate responses (n = 1), and surveys completed twice by the same person (n = 18) were removed, 632 pre survey and 373 post survey responses (59% of those who completed the pre survey) remained. It is noteworthy that due to the nature of data collection, available information around MECC trainers (changes pre and post training and fidelity to cascade of MECC training) was dependent on their data and the pre and post survey data of their trainees as entered into the MECC Gateway application. For core MECC training, surveys received an 87% response rate pre survey, 58% response rate post survey. For TtT, surveys received a 97% response rate pre survey and a 72% response rate post survey. As an incentive to complete both surveys, a certificate was only provided to attendees if they completed the post survey evaluation.

##### Materials

The regional evaluation surveys were created by the MECC regional at scale coordinator to evaluate the impact of the MECC TtT provided by the principal trainer. Thus, the questions were developed without input from the research team. Four questions were comparable across pre and post surveys (see Supplementary Material 1 for all survey questions). One related to confidence, which differed pre (‘How confident do you feel about having a training session on MECC?’) and post (‘How confident do you feel in delivering MECC training?’), one to motivation (‘How important is it for you to deliver MECC training?’), and one to knowledge (‘How would you rate your level of awareness of MECC?’). All three questions were answerable on a Likert scale of 1-5. The fourth question provided a grid of twelve words and asked participants to circle as many as were applicable. Additionally, eight questions assessed characteristics of the trainer such as their enthusiasm, curiosity, and eye contact, again rated on a Likert scale of 1-5. The post survey evaluation also asked participants how many courses they intended to deliver in the next 12 months (Intention, free text) and how ready they were to deliver training (Readiness, Likert 1-5).

### Data analysis

#### Differences pre and post training

Fidelity to receipt of treatment (training) of trainees was assessed using changes in (self-reported) confidence, motivation, and knowledge pre and post training. Additionally, changes in the frequency of each word in the presented grid that was highlighted could also be tested. As these words were unbalanced in terms of positive and negative words (eight suggested positive feelings and four more negative feelings), each word was analysed separately. After testing for normality assumptions (see Supplementary Material 2), a Willcoxon signed rank test was applied to all of the three variables. Chi-squared tests were applied to test the influence of mode of delivery (online or face to face) on the changes pre and post survey in Motivation, Confidence, and Knowledge. To analyse the question containing the word grid, the use of each word was broken down into ‘selected’ and ‘not selected’. An exact McNemar’s test was applied to each of the twelve words to assess for significant differences in the selection of each word pre and post training.

Existing literature assessing changes in confidence, motivation, or knowledge after receipt of MECC training does not report an effect size or sufficient data to calculate an effect size, thus a power analysis based on the average effect size across existing studies was not possible. However, one study reported on behavioural determinants [10]. A Gpower [42] power calculation estimated a minimum total sample size of 181 to detect a significant effect (matched pairs t test, two tailed, p ≤ 0.05), when the lowest of the reported effect sizes (0.27) was considered (the most conservative estimation). Therefore, the available sample size was deemed as sufficient to statistically detect effects on confidence, motivation, and knowledge.

Additionally, differences in the baseline characteristics of those who did (n = 373) and did not (n = 259) complete post evaluation measures were tested for statistical significance. Again, the data did not meet normality assumptions (see Supplementary Material 2), thus Mann-Whitney U tests were applied to test baseline differences in completers and non-completers. Completers scored significantly higher in baseline Knowledge (U = 42578.50, z = −2.64, p = .008) than non-completers, despite both medians being equal (Median = 3). There were no significant differences in baseline Confidence (U = 46672.00, z = -.76, p = .449) or Motivation (U = 45979.50, z = −1.09, p = .274) scores between completers and non-completers, for both variables and groups within each variable the median was equal (Median = 4).

### Fidelity to training cascade

To preserve the anonymity of participant secondary data, it was not possible to link pre/post TtT survey data with information concerning training cascade for each trainer. Instead, descriptive statistics were calculated to explore the number of trainers who had delivered training, the number of sessions they delivered, and the number of people they had trained. Descriptive statistics were used to examine self-reported fidelity to training cascade in terms of adherence and dosage. Adherence measures the adequate delivery of the key components of the intervention; in other words, delivery of the intervention as it was designed. Dosage describes the amount of the intervention delivered. Namely, fidelity related to study design was measured using the frequency of TtT sessions delivered by the principal trainer, although the actual dose (duration) and content of the sessions was not recorded. Training providers can be viewed as the principal trainer or trainers that have attended TtT training, although only self-reported data on the change in the knowledge and confidence of trainers is available to assess this area, through pre and post evaluations.

### Predicting intentions to cascade

Sample size did not reach adequate statistical power to apply an ordinal regression with Readiness as the dependent variable (see Supplementary Material 2). Thus, multiple linear regression was applied using the ‘Enter’ method in SPSS [43] with Intention as the dependent variable (see Supplementary Material 2 for assumptions testing), which required a sample size of 98 to detect a small effect size (.20) when a power analysis was conducted (linear multiple regression: fixed model, R^2^ deviation from zero, two tailed, p < 0.05, power .95). Normal distribution of standardised residuals was skewed by ten outliers of higher values (Intentions of between 8-10 sessions, see Supplementary Material 2 for details). However, as it was important that the model represented individuals who intended to deliver a high number of sessions, the outliers were retained for the analysis. A sensitivity analysis was conducted which removed the ten outliers (regression coefficients and standardised errors are presented in Supplementary Material 3).

#### 2. Analysis of primary and secondary qualitative data

##### Participants

Interview participants (n = 21) were either eligible to receive MECC TtT training and had (n = 13) or had not (n = 6) received it or were principal trainers within the NENC region (n = 2). To ensure a representative sample in terms of the variation of current MECC training delivery, participants were purposively sampled. The Regional Delivering MECC at Scale Coordinator sent a recruitment advert to eligible participants within their existing networks and participants were asked to fill out an expression of interest form with basic demographic information including sector (e.g local authority, the third sector, and healthcare settings), nature of attendance (invited to attend or self-referred), and history of attendance (attended or not attended), and a variation of participants were selected to participate.

Target sample size was guided by information power [44] and defined apriori; given that the aim was specific, sampling was purposive, analysis was pragmatic and applied an established theoretical framework, and rapport was built with participants by an experienced qualitative researcher, the total target sample size was 20.

##### Materials

There were two topic guides for those who had and had not attended MECC TtT training. The former topic guide was amended for participants who were principal trainers (see the supplementary material of the in-depth qualitative paper [39]). Topic guides were informed using the TDF [35, 45] and explored barriers and facilitators to accessing the MECC TtT programme and cascading as well as the acceptability of the TtT model. Strategies to increase training cascade gathered from existing literature were extracted using a triangulation process. To explore participants’ opinions, strategies cited by at least two studies focused and not focused on MECC each were proposed to participants at the end of the interview. The proposed strategies were refresher training [20, 27, 46–48], which was the most frequently suggested, and peer support groups [18, 27, 46, 49]. Additionally, responses from the free text comments of the post survey were used to inform the findings (Supplementary Material 1, more details in the in-depth qualitative paper: [39]).

##### Procedure

Interviews were conducted online via Teams (n = 18) or in person at the organisation of participants for non-NHS staff (n = 2) or *(blinded for peer review)* campus (n = 1). All interviews were conducted by the lead author (*blinded for peer review)* who is experienced in conducting interviews relating to MECC at the time of interviews, and was already known as a PhD student and research assistant in MECC research to some but not all participants. Where the participant was unknown to the interviewer, an effort was made to build rapport prior to interviews and the interviewer introduced themselves as a research assistant on the project. After providing participants with the information sheet and gaining written informed consent, the relevant topic guide was flexibly applied during a semi-structured interview. Participants were sent the debrief sheet on completion of the interview. A think aloud protocol was applied [50] with the triangulated strategies described above, as participants were presented with one strategy at a time and asked to rate each one in accordance with the APEASE criteria (as described in the professional contribution section) [34] whilst providing their thoughts verbally in real time. However, during interviews, there was often not enough time remaining to conduct the think aloud protocol and thus it was not conducted consistently. Interviews were digitally recorded and transcribed verbatim via a transcription service. Interviews lasted between 18 and 88 minutes.

##### Data analysis

Qualitative data was coded according to TDF domains [35], although interview and survey data were coded separately with survey data coding as secondary. Coding of TDF domains was applied using the target behaviour of cascading MECC training, although free text comments could be more difficult to code due to a lack of context.

Comments were not coded if they were too vague to code to a specific TDF domain, related to MECC delivery, or it was not clear whether the comment related to MECC delivery or MECC training delivery. However, comments related to the MECC training that could be related to the subsequent delivery of MECC training were coded (e.g comments around the need for more information on how to conduct a MECC conversation were coded as this may influence how confident trainers feel to deliver MECC training or how useful they perceive MECC training to be to their staff). To ensure inter-rater reliability, 10% of both the interviews and qualitative post survey comments were analysed by an independent rater (*blinded for peer review*) and disagreements were resolved through discussion between all three raters such that one independent rater resolved conflicts between the other two raters. Qualitative analysis software NVivo was utilised [51]. A coding system was used to label transcripts of principal trainers (PT), those who had attended the MECC TtT training (AT), and those who had not attended the MECC TtT training (NA), respectively. In accordance with open data procedures, fully anonymised transcripts are available via the UK Data service [52].

#### 3. and 4. Content analysis of the TtT programme and SBA

##### Materials

TtT programme content and resources provided to MECC trainers were gathered from the regional principal trainer and Regional Delivering MECC at Scale Coordinator. To provide a more comprehensive analysis of the training and improve inter rater reliability (explained below), authors (*blinded for peer review*) observed a MECC TtT training session on the 11^th^ November 2023. The training was also audio recorded, which was placed at the front of the room and captured the full training session. A combination of access to the slides and observation of the training session ensured all relevant written and oral BCTs were captured.

To facilitate the mapping of TDF domains onto BCTs, an existing mapping tool was utilised (https://theoryandtechniquetool.humanbehaviourchange.org/tool). The tool identifies links between the two through convergence of findings from a literature review and Delphi consensus study between behaviour change experts. It is noteworthy that many links have not yet been investigated, thus absence of a link does not necessarily indicate that a TDF and BCT are not linked.

##### Data analysis

The MECC TtT programme was analysed for the presence of BCTs using the hierarchically clustered taxonomy [36] and intervention functions utilised [34] by one author (*blinded for peer review*). To ensure inter rater reliability of coding, a second author (*blinded for peer review*) independently coded the slides for BCTs and intervention functions utilised. Inter rater reliability was calculated using Kappa, using the conservative parameters set by Altman, identifying agreement between raters to be poor (k = .15). Thus, a third author (*blinded for peer review*) live coded the TtT training during observation of training delivery. This time, inter rater reliability between the lead author (*blinded for peer review*) and the independent rater (*blinded for peer review*) was good (k = .64). Disagreements were resolved through discussion between all coders (*blinded for peer review*) to determine final coding.

Qualitative data from interviews and post-survey free text comments were triangulated to inform key TDF domains for the SBA. Namely, key TDF domains were identified by triangulating the following; 1) frequency of transcripts that TDF was coded, 2) frequency of times that TDF was coded in post survey comments, and 3) elaboration of TDF domain (number of sub-themes). Specifically, TDF domains were ranked based on each criterion, and the mean ranking was calculated across the three criteria to inform on which TDF domains were identified as key domains. Key TDF domains were then mapped onto the identified BCTs and intervention functions within the MECC TtT programme to identify whether current training helps address these barriers and where there are missed opportunities.

### Triangulation of data

Qualitative and quantitative data were triangulated both through quantitising (comparing the key TDF domains identified through calculating frequency and depth of use and ranking them with the findings from the comparisons between pre and post surveys and the multiple regression) and qualitising (using narrative description of the findings to triangulate with the qualitative data) after all data was collected [53]. Combining all data informs on a holistic perspective of the challenges associated with the MECC TtT model and how its implementation may be improved.

### Findings

#### 1. Secondary analysis of quantitative data

Descriptive statistics for the quantitative data are presented in Tables 1 and 2. The questions regarding characteristics of the trainer scored highly (Means ranged between 4.65 to 4.84), with no participants selecting ‘1’ for any. Responses to readiness and intentions (range: 0-10) were on average lower and with a wider range of responses. Concerning the pre and post variables (Knowledge, Confidence, and Motivation), all responses pre and post utilised the maximum possible range of 1 to 5, aside from post survey Knowledge, of which no participants selected 1. Pre training, participants showed a lower mean Knowledge score (M = 2.72) compared to Confidence (M = 3.87) and Motivation (M = 4.1).

**Table 1:** Descriptive statistics for the quantitative data. SD = standard deviation. The questions that informed each variable are provided in Supplementary Material 1.

|  | Mean | SD |
| --- | --- | --- |
| **Non completers (n = 259)** |  |  |
| Confidence | 3.9 | 1.04 |
| Motivation | 4.0 | 1.11 |
| Knowledge | 2.5 | 1.08 |
| **Completers pre (n = 373)** |  |  |
| Confidence | 3.87 | .93 |
| Motivation | 4.1 | .98 |
| Knowledge | 2.72 | 1.09 |
| **Completers post (n = 373)** |  |  |
| Confidence | 3.84 | .84 |
| Motivation | 4.1 | .94 |
| Knowledge | 4.1 | .68 |
| Intentions | 3.04 | 1.88 |
| Readiness | 3.31 | .94 |
| Trainer: listened | 4.83 | .43 |
| Trainer: positive response | 4.84 | .42 |
| Trainer: positive language | 4.78 | .48 |
| Trainer: enthusiastic | 4.74 | .53 |
| Trainer: eye contact | 4.82 | .45 |
| Trainer: used names | 4.72 | .62 |
| Trainer: learning into practice | 4.65 | .66 |
| Trainer: curiosity in topic | 4.73 | .54 |

**Table 2:** Frequency that each proposed word was selected at pre and post survey (n = 373), and the corresponding result of the exact McNemar’s test. There was a significant increase in the selection of words ‘knowledgeable’, ‘confident’, ‘energised’, ‘competent’, ‘enthused’ and significant decreases in the selected of words ‘uncertain’, ‘apprehensive’, and ‘focused’. There was no significant differences in the use of words ‘confused’, ‘frustrated’, ‘motivated’, or ‘positive’.

| Word | Pre | Post | *p* |
| --- | --- | --- | --- |
| Confident | 61 | 187 | < .001 |
| Uncertain | 68 | 13 | < .001 |
| Energised | 24 | 52 | < .001 |
| Competent | 31 | 94 | < .001 |
| Apprehensive | 33 | 16 | .009 |
| Enthused | 70 | 94 | .031 |
| Knowledgeable | 5 | 111 | < .001 |
| Confused | 5 | 0 | .063 |
| Frustrated | 0 | 1 | 1 |
| Motivated | 93 | 91 | .925 |
| Positive | 136 | 129 | .630 |
| Focused | 47 | 25 | .005 |

##### Differences pre and post training

Wilcoxon signed-rank tests identified no significant differences in Confidence (Z = -.45, p = .650) or Motivation (Z = −1.03, p = .302), but a significant improvement in Knowledge (Z = - 15.17, p < .001). Indeed, the median for both Confidence and Motivation remained at 4 pre and post training, whilst the median for Knowledge increased from 3 to 4 on a scale of 1 to 5. There were no significant differences between those who attended training online or face to face, that is for both modes of delivery there was no change in Motivation (χ(1) = 4.12, p = .846) or Confidence (χ(1) = 4.55, p = .715), and most participants improved by one scale point for Knowledge (χ(1) = 1.67, p = .898), with differences following a normal distribution around this mode.

##### Fidelity to training cascade

In total, 1051 people received core MECC training, and 705 attended TtT training. Only 4.4% had delivered MECC training (n = 31), according to the registered trainer on pre core MECC training surveys. Of those that had cascaded, only 41.9 % (n = 13) had delivered more than one training session and 22.5% (n = 7) had delivered more than two. Number of MECC training sessions delivered ranged from 1 to 41 (by the principal trainer, M = 4.39, SD = 8.11), and the number of people each trainer trained ranged from 1 to 170 (again, the largest number delivered to was by the principal trainer, M = 26.42, SD = 45.06). Thus, when removing the principal trainer from analysis, the maximum number of MECC training sessions delivered was 21 (M = 3.17, SD = 4.50), the maximum number of people each trainer trained was 154 (M = 21.63, SD = 36.96).

##### Predicting intentions to cascade

The effect size of the prediction of Knowledge, Motivation, Confidence, and mode of delivery on number of sessions individuals intended to deliver was very small [54] but significant (F(4,368) = 19.637, p < .001), with an R^2^ of 17.6% and an adjusted R^2^ of 16.7%. Neither mode of delivery nor Knowledge were significant predictors, although Motivation and Confidence were (see Table 3 for regression coefficients and standardised errors).

**Table 3:** Regression coefficients and standardised errors for the multiple linear regression analysis to predict number of MECC training sessions intended to deliver. B = unstandardised regression coefficient, CI = confidence interval at α = .05, SE B = standard error of the coefficient, β = standardised coefficient.

| Independent variable | $B$ | 95% CI for $B$ | | SE $B$ | $\beta$ |
| --- | --- | --- | --- | --- | --- |
|  |  | LL | UL |  |  |
| Mode of delivery | .052 | -.428 | .532 | .244 | .010 |
| Knowledge (post) | .1444 | -.181 | .469 | .165 | .052 |
| Motivation (post) | .576 | .344 | .809 | .118 | .288 |
| Confidence (post) | .333 | .077 | .589 | .130 | .150 |

Sensitivity analyses with the removal of participants who intended to deliver > 8 sessions resulted in the effect size remaining small (19.9% with an adjusted R2 of 19%) but significant (F(4,353)= 21.970, p < .001), with the same significant predictors as the original model (see Supplementary Material 3).

#### 2. Analysis of primary and secondary qualitative data

Table 4 displays the prioritisation of TDF domains alongside key barriers and facilitators within each domain, although the following quote aptly summarises the most fundamental barriers to training cascade that participants described.

> *‘So the problem we’ve had, which I’m guessing, and I know it’s not unique, not that, I’m guessing it’s not unique, people go away, back to their workplaces, and they aren’t supported to deliver their training when they get there. So we had lots of feedback around, training was very very useful and enjoyable, but I don’t have the capacity when I get back into my team, either the managers not supportive of the extra time it’s going to take, or we didn’t actually realize what we were signing up to when we came on your programme, even though the clue is kind of in the title a bit, or actually, I’ve really enjoyed it, but I don’t feel confident or comfortable delivering the MECC training. That’s not, that’s not me, and that’s not what I want to do. So I would say, of a cohort of about, I think we managed in the end to get about 74 people through MECC train the trainer. So we were doing it once a month on average between September and April. Of those 74, I would say we’re lucky to have maybe 10% of those staff delivering core MECC routinely.’* (AP13)

**Table 4:**
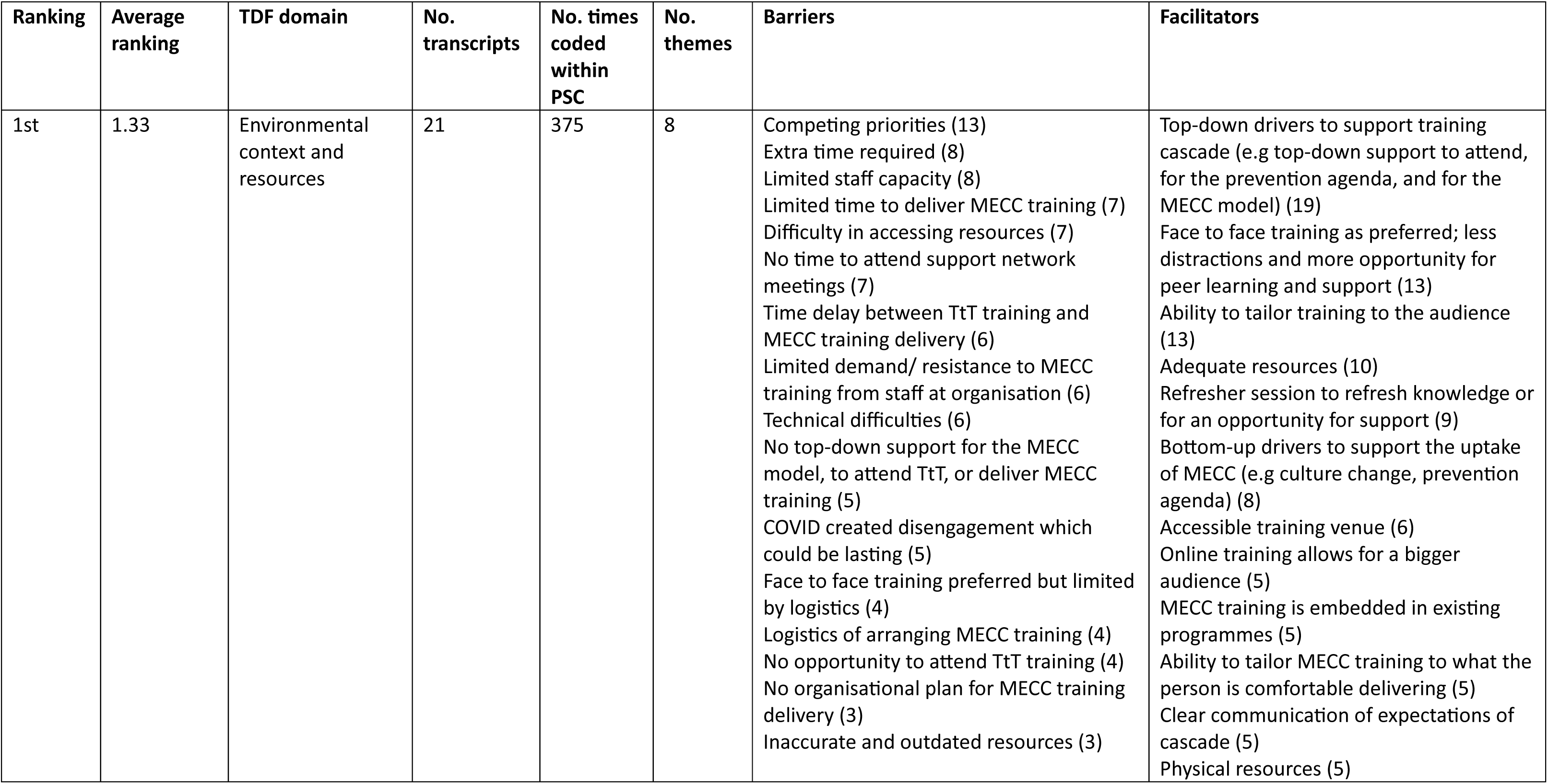
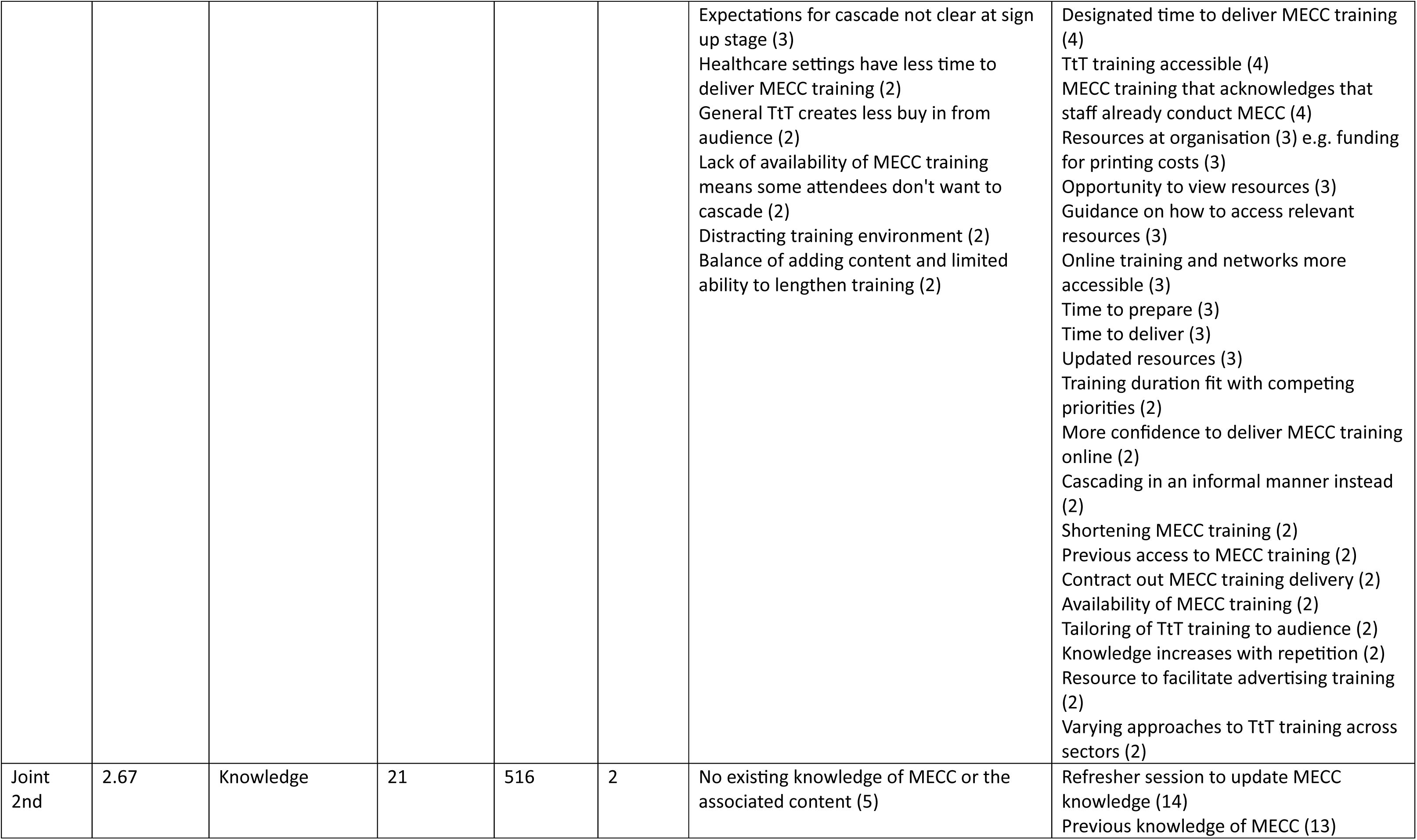
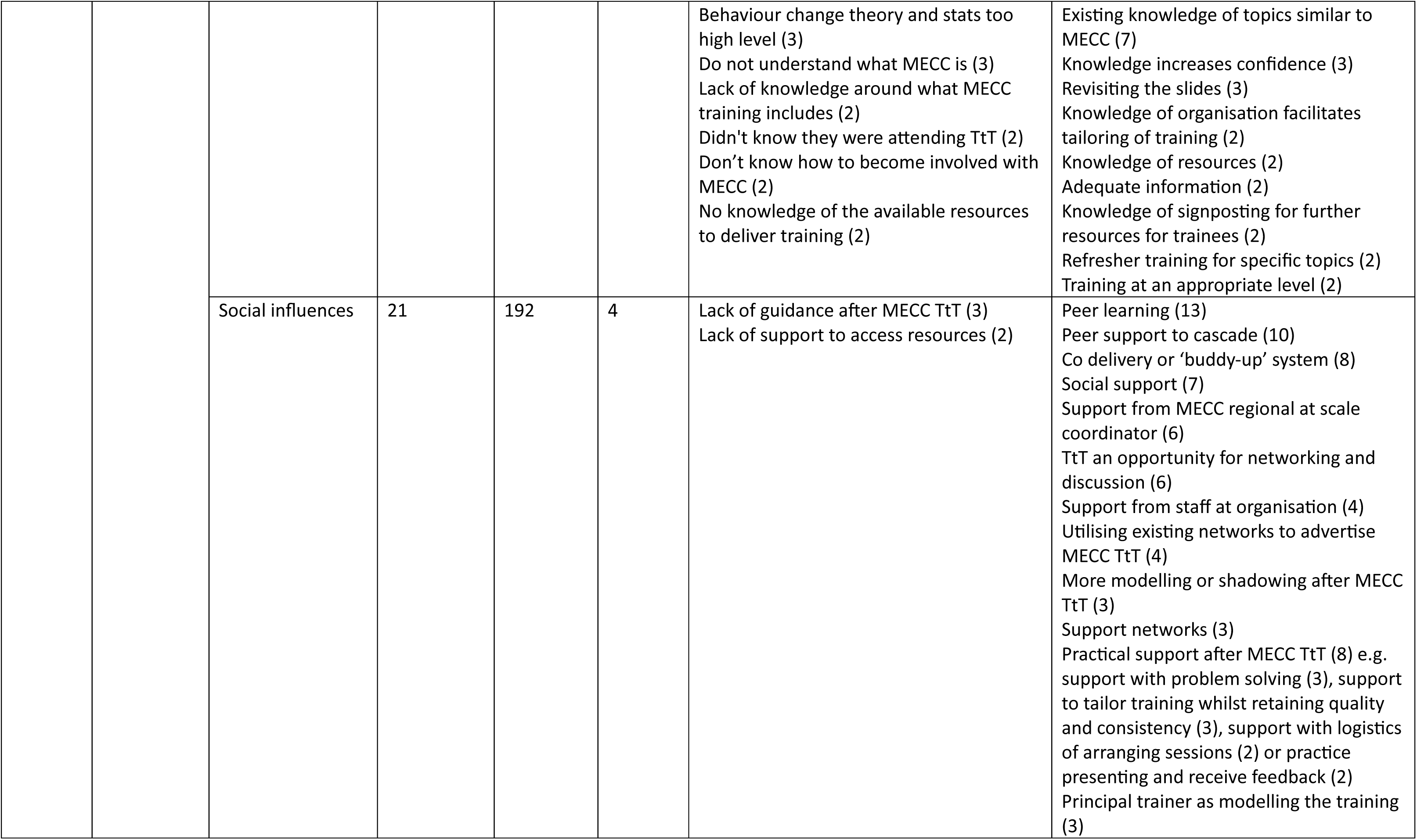
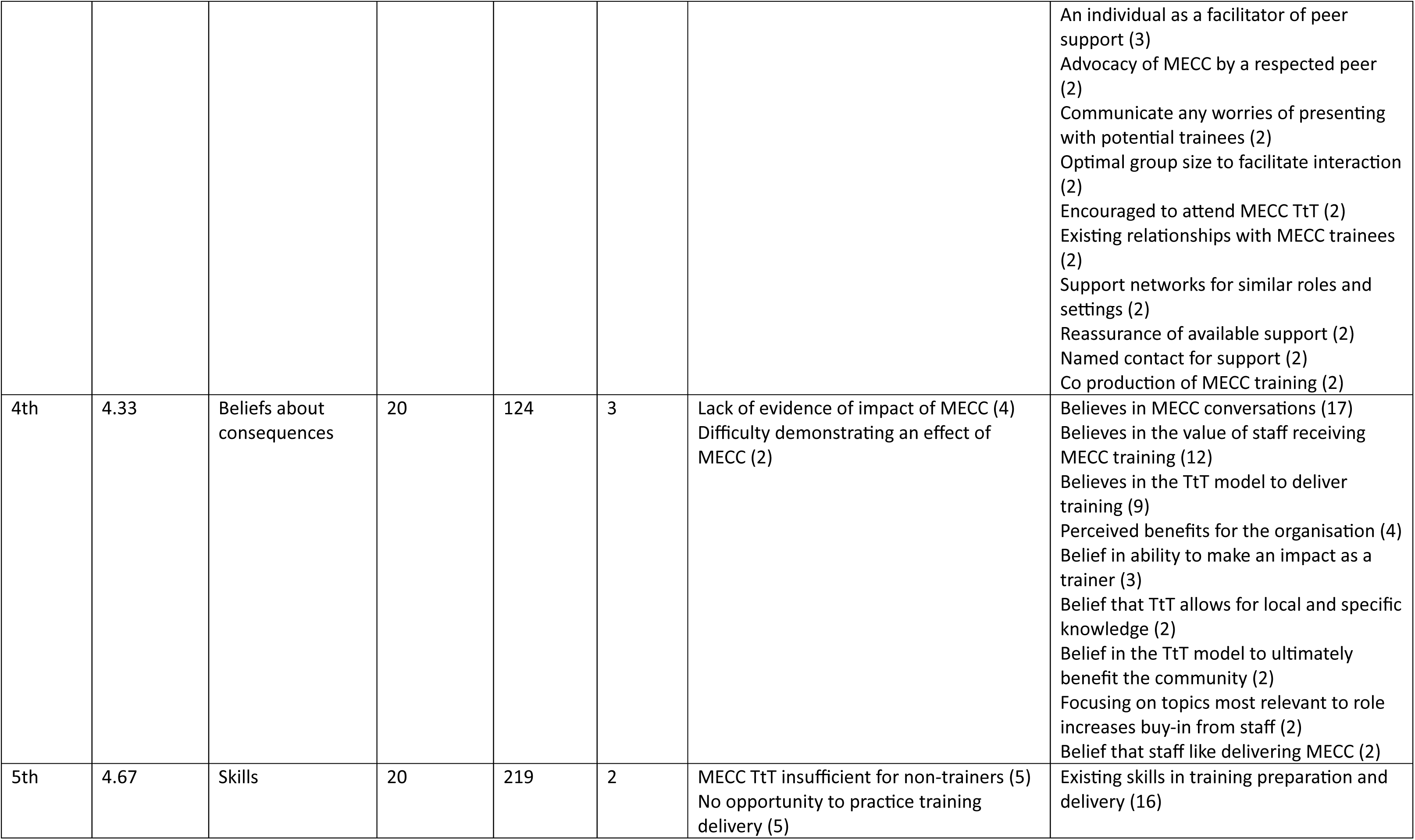
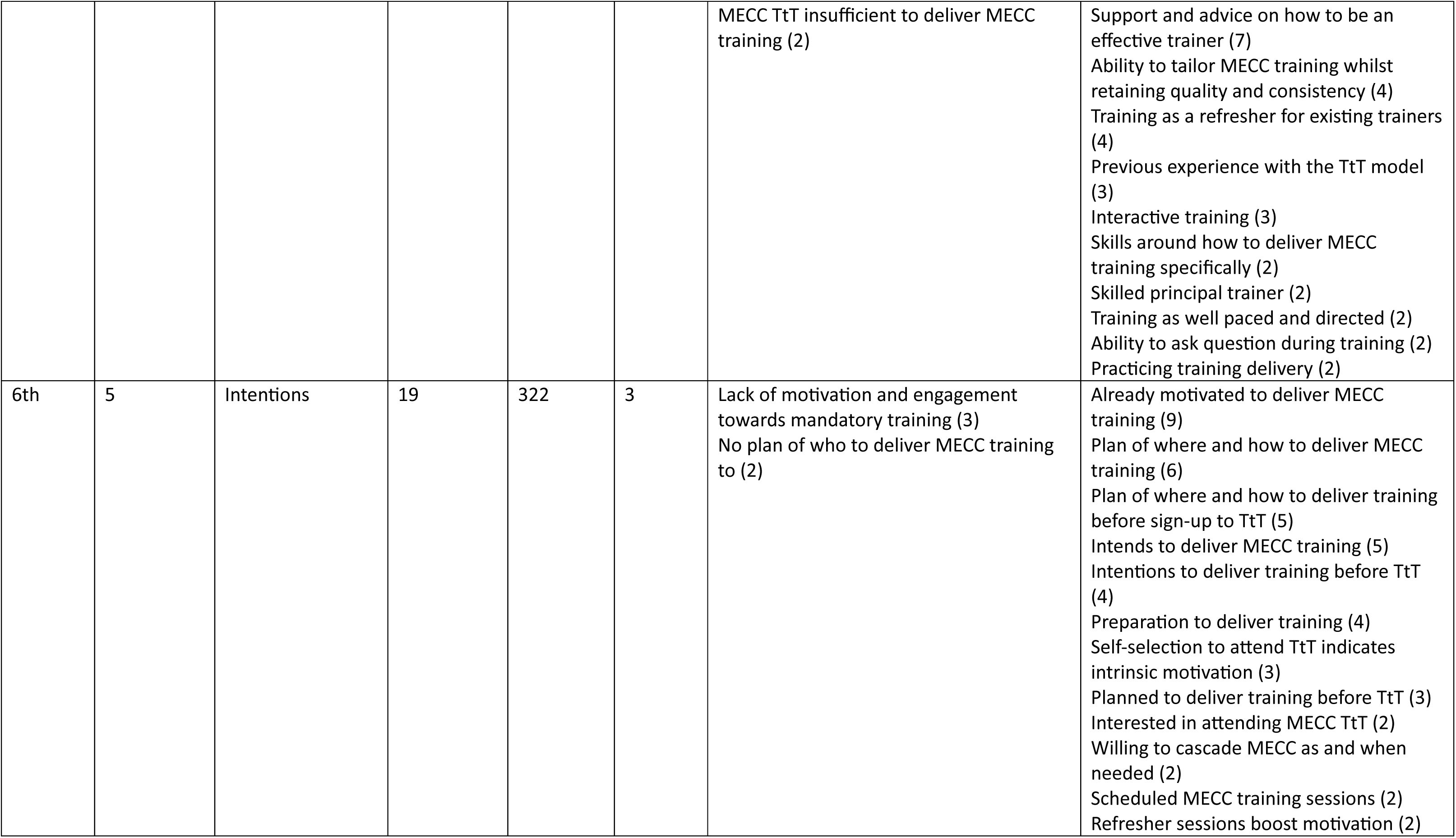
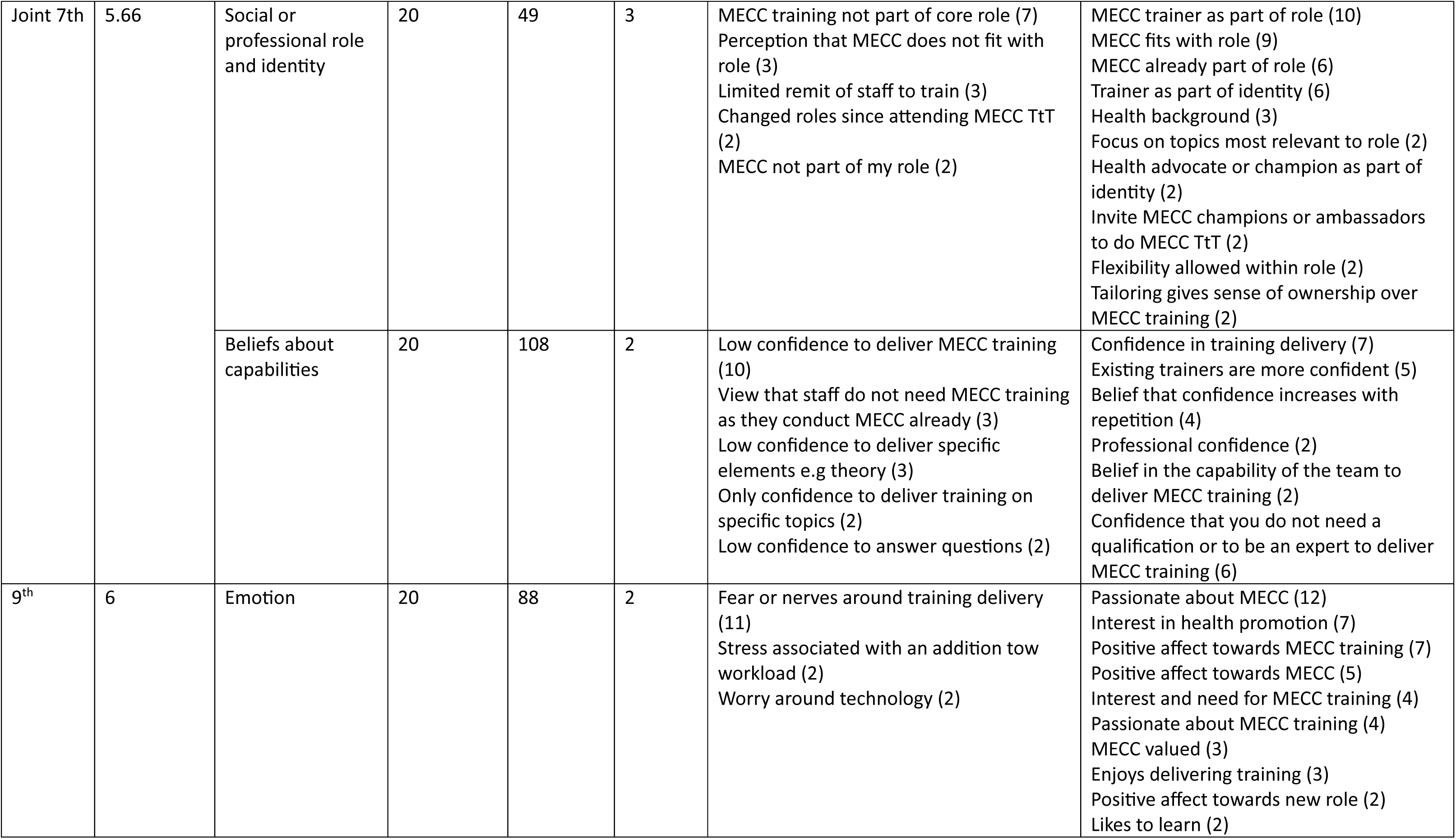
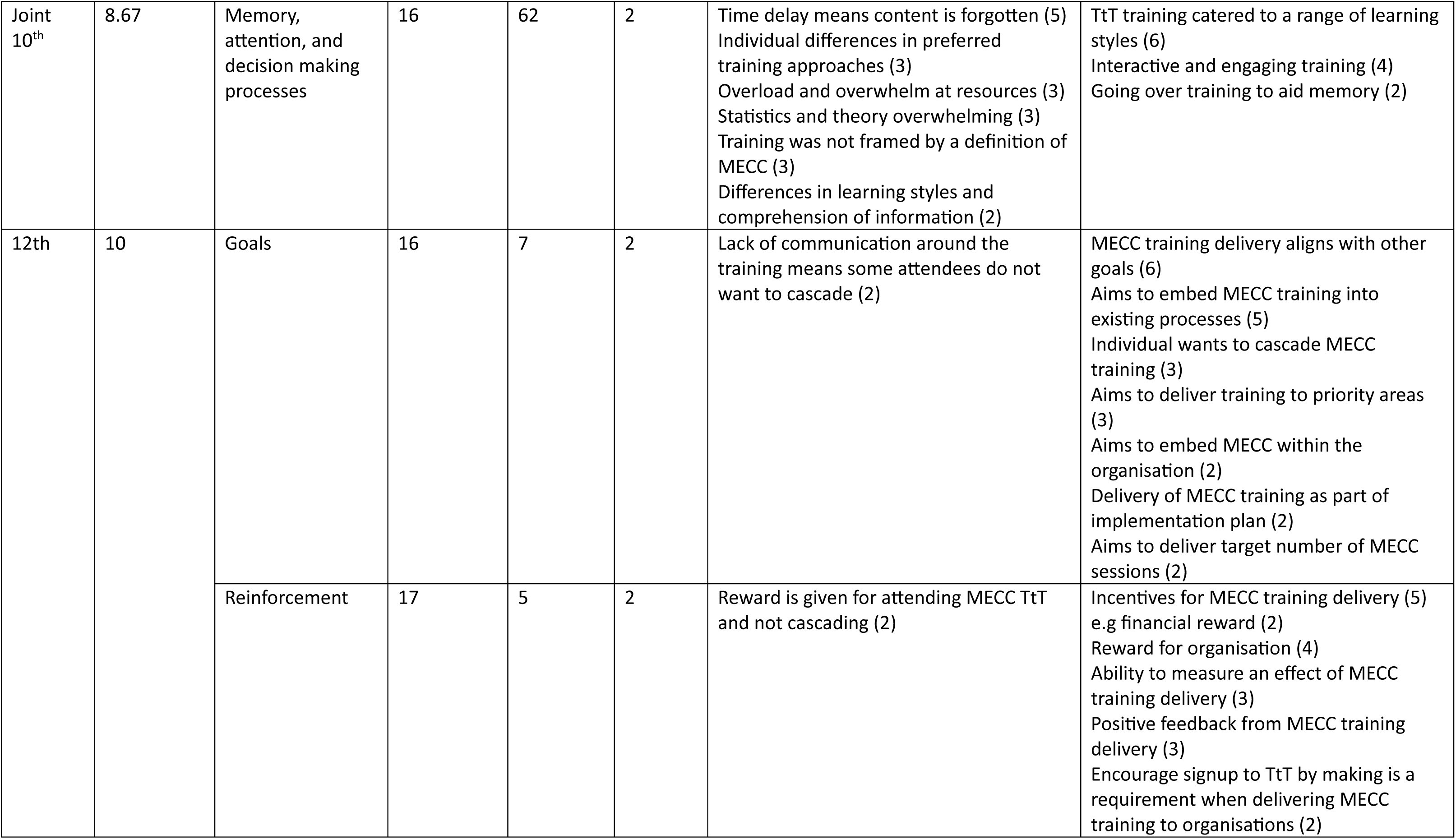
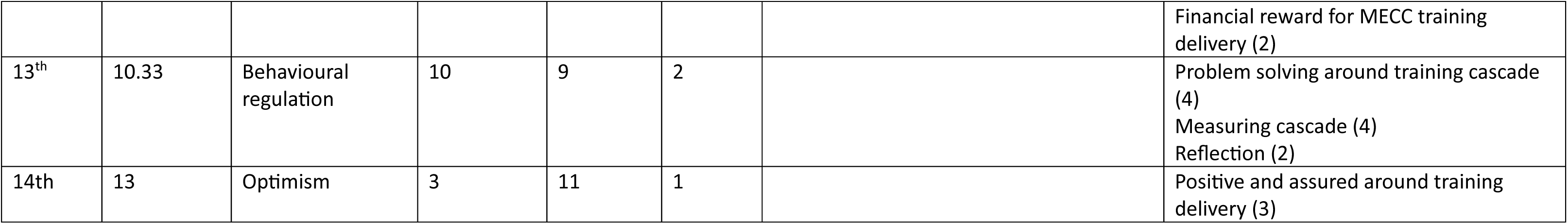
Prioritisation of TDF domains according to number of transcripts each was coded within, number of times the domain was coded within the post-survey comments (PSC), and elaboration of each domain (number of themes within each). Barriers and facilitators within each domain are also presented, with the frequency of transcripts each was cited in displayed in brackets. Only barriers and facilitators identified by more than one participant are included.

##### Barriers and facilitators to cascade

Six key TDF domains were identified; Environmental Context and Resources, Knowledge, Social Influences, Beliefs about Consequences, Skills, and Intentions. Particularly, the domain Environmental Context and Resources was the most important domain associated with the barriers and facilitators to cascade of MECC training. The most frequently cited barriers within this domain included 1) lack of time due to limited capacity and competing priorities, 2) lack of top-down support to cascade MECC training, 3) resistance of staff towards MECC training, 4) difficulties with accessing and using technology to prepare and deliver MECC training, 5) a time delay between attending the MECC TtT programme and cascading MECC training, 6) interruption of the COVID pandemic to MECC implementation plans, 7) challenges with the logistics of organising MECC training, and 8) a lack of access to the MECC TtT programme to interested individuals. Meanwhile, the most frequent facilitators relating to Environmental Context and Resources were 1) top-down support for cascade including support of the MECC model and to attend, prepare for, and deliver MECC training, 2) adequate resources with an opportunity to tailor MECC training to the audience and what the MECC trainer was comfortable in delivering, 3) refresher sessions, 4) bottom-up drivers for the uptake of MECC including culture change, 5) clear communication of expectations of cascade prior to the MECC TtT programme, 6) a designated time or existing training programme to deliver MECC training within, 7) physical resources that MECC trainees could take away from the MECC TtT programme such as a training booklet, and 8) training that was accessible both in terms of location and accessibility requirements.

Other important barriers to cascade centered around characteristics of the trainers, including fear and worry around training delivery (Emotion), low confidence in training delivery (Beliefs about Capabilities), MECC training delivery as not part of an individual’s core role (Social or Professional Role and Identity), no pre-existing knowledge related to MECC (Knowledge), and no intention to deliver MECC training after receipt of MECC TtT training (intentions). Many of these were compounded by barriers relating to the MECC TtT programme, including the training as insufficient for those who had not delivered training before (Skills), no opportunity within the training to practice training delivery (Skills), a lack of evidence of impact of MECC (Beliefs about Consequences), and a lack of guidance after MECC including to access resources (Social Influences).

Frequently cited facilitators under other TDF domains that were specific to the MECC TtT programme included peer learning or support such as through a buddy-up system for training delivery or support networks (Social Influences), confidence in training delivery which was increased for existing trainers or with repetition of training delivery (Beliefs about Capabilities), advice on how to deliver training (Skills), support from a key contact such as the Regional Delivering MECC at Scale coordinator who could act as a facilitator of peer support (Social Influences), practical support such as technical advice (Social Influences), incentives for training delivery (Beliefs about Consequences), training that catered to a range of learning styles (Memory, Attention, and Decision Making Processes), and interactive and engaging training (Memory, Attention, and Decision Making Processes). Other important facilitators included previous experience in training preparation and delivery (Skills), previous knowledge of MECC or related topics and the opportunity to update knowledge after training (Knowledge), belief in the impact of staff receiving MECC training and ultimately delivering MECC conversations (Beliefs about Consequences), MECC and to a greater extent delivering MECC training as seen to be part of or relevant to the individual’s role (Social or Professional Role and Identity), motivation and plans to deliver training before attending the MECC TtT programme (Intentions), a passion, interest, and positive affect towards MECC and delivering MECC training (Emotion), support for the TtT model to deliver training (Beliefs about Consequences), an ability to tailor MECC training whilst retaining the fundamental elements (Skills), cascade as aligning with other goals such as embedding MECC within the organisation (Goals), problem solving around how to cascade training (Behavioural regulation), and the ability to measure cascade (Behavioural regulation).

##### Relative influence of barriers and facilitators to cascade

To better understand which of the many barriers and facilitators were instrumental in determining cascade of training, Table 5 maps the characteristics of all participants who had attended the MECC TtT programme against whether they had cascaded or planned to cascade MECC training. As Table 5 demonstrates, an existing passion for MECC was not necessary or sufficient to determine cascade and relying on passion with no training experience or if training delivery was not part of an individual’s role meant that cascade was prevented if individuals faced personal challenges. On the other hand, top-down support and existing training experience, particularly if training was part of an individual’s role, were instrumental in determining cascade.

**Table 5:**
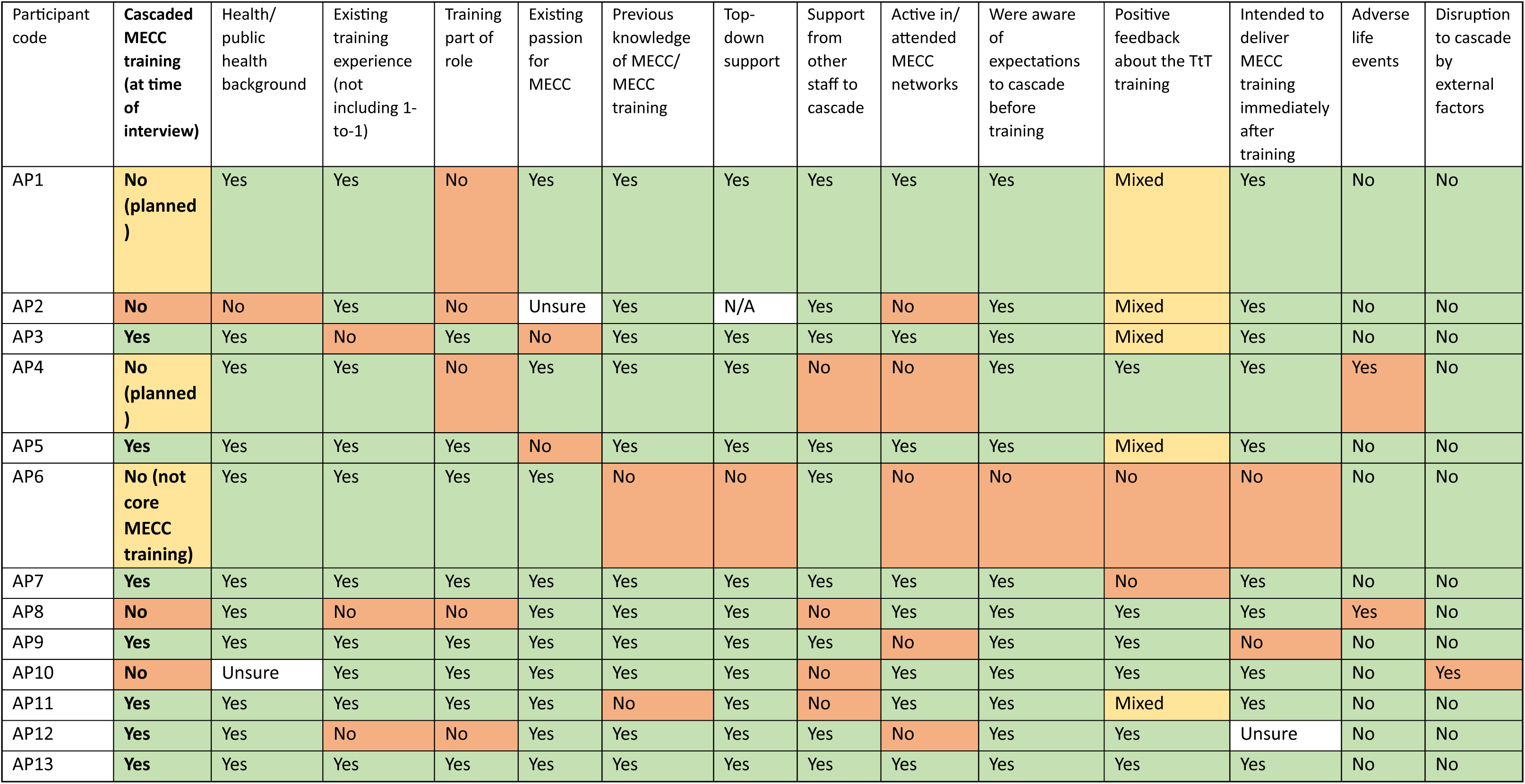
profile of each TtT attendee interviewed, according to important identified characteristics during analysis.

Unsuccessful cascade, despite training being part of participants’ role, could be attributed to external barriers outside of their control. Only one participant that had successfully cascaded MECC training had no training experience and MECC training delivery was not part of their role, however they possessed other key facilitators including top-down support and support from staff which was sufficient to overcome this barrier. There were however differences in degree of top-down support; whilst some participants noted that their leadership and management approved of them attending the MECC TtT programme, others took a more active and facilitating role such as attending the training with them, allowing specified time for practice and delivery, and encouraging cascade to fulfil a MECC implementation plan, with the latter indicating a higher chance of successful cascade. Whilst many of those successful in cascading were involved in MECC networks, the interview findings showed that those who had not yet cascaded were less likely to attend as they felt *‘embarrassed’* or like the meeting was not relevant to them. However, it is also clear that a single factor did not appear to determine successful cascade, rather it was a combination of factors, some more influential than others. It is noteworthy that some participants (AP1, AP10) had provided significant input into the planning and preparation phase, so whilst not yet cascaded the training, there was indication that the individual would later be successful in cascading MECC training.

#### 3. Content analysis of the TtT programme

Properties of the training programme are shown in Table 6. Seven BCTs (see Table 7) and four intervention functions (see Table 8) were identified within the TtT training.

**Table 6:** Tidier checklist of the MECC TtT training.

|  |  |
| --- | --- |
| Brief name | MECC train the trainer training |
| Why | Resources are limited, thus a cascade model aims to provide MECC training and ultimately MECC delivery to as many individuals in the region as possible. Only one principal trainer is required if attendees deliver their own MECC training sessions (cascade) after TtT training |
| What (material) | Attendees are first shown the core MECC slide deck and videos and case studies that are tailored to the audience. Attendees are shown the MECC gateway signposting tool and where to access training resources (NHS Futures website), and how to use the ICS to arrange MECC training. Tips and advice on training delivery are provided advised with a 'diamond nine' group task to prioritise key behaviours and skills of trainers. Attendees are instructed to tailor their training to their audience, plan and practice, record their training delivery on the ICS app, join support networks, and deliver at least 4 sessions annually, with the first one delivered in three months. |
| What (procedures) | The first two hours of the session runs through core MECC training as it is delivered (through a didactic approach). Attendees are provided with a comfort break. The final 45 minutes focuses on how to deliver MECC training. |
| Who provided | The principal trainer, who is highly experienced in nursing and training/education. |
| How (mode of delivery) | Most training sessions are delivered face to face, with a general guide of between five to twelve attendees to facilitate group discussion. |
| Where | Venues are selected by the organiser (e.g a member of staff from healthcare or local authority that arranges attendees and requests the principal trainer), thus are local to them (e.g a community centre). |
| When and how much | One, half day (3.5 hour) training session. |
| Tailoring | The topics discussed in the core MECC training section are selected based on the audience, so that they are relevant to them but not already part of their expertise e.g for those working in smoking cessation, two topics would be selected that were considered not within their current expertise such as mental health and diet. There is no tailoring of the train the trainer section. |
| Modification | The programme is continuously modified in response to feedback from attendees (e.g using different theories, different illustrations of theories, adding new topics), although the train the trainer section remains mostly stable across time. |
| How well (planned) | Trainees are encouraged to attend support networks including the MECC trainers forum which hosts online meetings. Trainees are also encouraged to record the number of training sessions delivered and number delivered to via the ICS app, although due to logistical challenges and an organisations monitoring cascade internally the regional data is not comprehensive. |
| How well (actual) | Only 4.4% of those who have received TtT training have delivered training. Of these, only 13 delivered more than one training session and 7 delivered more |
|  | than two. The principal trainer has delivered the most MECC training sessions to the largest number of trainees. |

**Table 7:** Seized and missed opportunities according to the congruence between BCTs utilised within MECC TtT training and the key TDF domains identified. TDF domains highlighted in bold are the key six domains identified from content analysis of barriers and facilitators. If TDF domains were ranked equally, both have been provided with the higher ranking (e.g 1 if joint first and second). *The integrated matrix maps BCTs onto TDF domains for links between them which can be accessed here: https://theoryandtechniquetool.humanbehaviourchange.org/tool. **Judgement of congruence is according to the number of key TDF domains that are linked with the BCT: low = none, medium = one, high = two or more.

| BCT | Example of BCT in the MECC TtT training | TDF domains (from integrated matrix*) | Domain Importance ranking | Theoretical congruence with TDF domains** |
| --- | --- | --- | --- | --- |
| Goal setting (behaviour) | Trainees are advised through a 'memorandum of understanding' to deliver at least 4 MECC sessions annually, first within 3 months | **Intentions**<br>Goals | 6<br>12 | Medium |
| Instruction on how to perform a behaviour | Discussion of the relevant skills needed to become a MECC trainer, advise on training delivery | **Knowledge**<br>**Skills**<br>Beliefs about capabilities | 2<br>5<br>7 | High |
| Self-monitoring of a behaviour | Trainees are advised through a 'memorandum of understanding' to use the ICS Training App to organise & deliver MECC training | Behavioural regulation | 13 | Low |
| Social support (unspecified) | Utilisation of group discussion | **Social influences** | 2 | Medium |
| Demonstration of the behaviour | Didactic training approach- principal trainer first delivers MECC training | Beliefs about capabilities | 7 | Low |
| Behavioural practice/rehearsal | Trainees are advised to 'prepare and rehearse' | **Skills**<br>Beliefs about capabilities | 5<br>7 | Medium |
| Adding objects to the environment | Trainees are provided with resources for training delivery including signposting resources | **Environmental context and resources** | 1 | Medium |

**Table 8:**
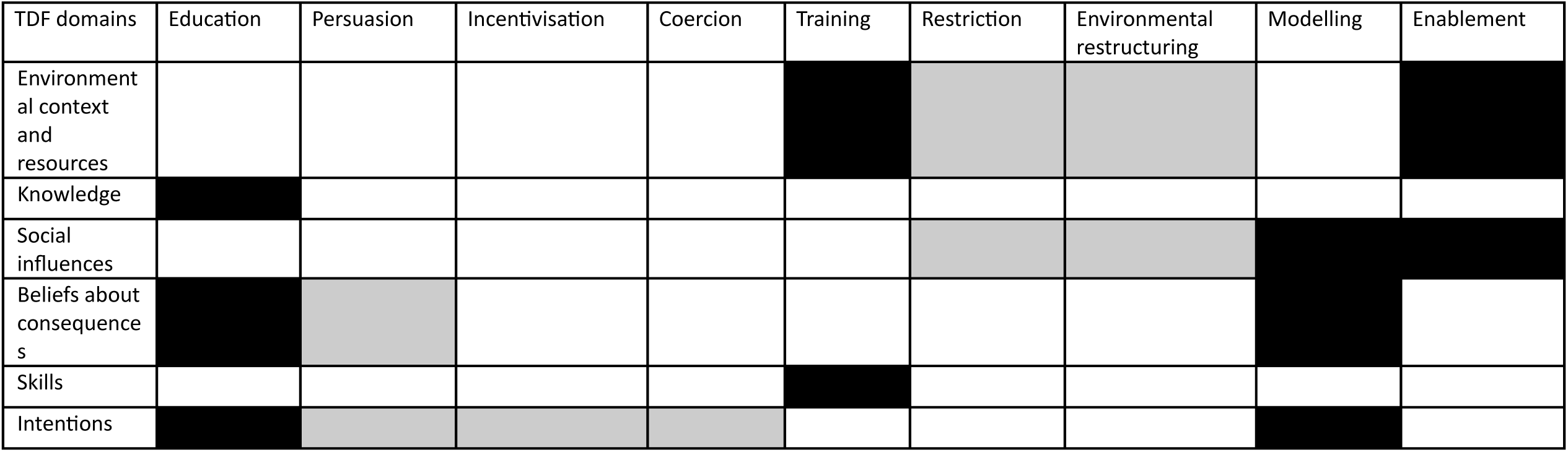
Opportunities missed and seized in terms of IFs against the key seven TDF domains, using mapping of links between TDF domains and IFs from Michie et al. (1). IFs define the columns. Black = seized opportunity, light grey = missed opportunity.

#### 4. Strategic Behavioural Analysis

The only included BCT within MECC TtT training that showed high congruence (acknowledged two or more key TDF domains) with the relevant TDF domains was instruction on how to perform a behaviour. Four addressed at least one key TDF domain (goal setting (behaviour), social support (unspecified), behavioural practice/rehearsal, and adding objects to the environment) and two addressed none (self-monitoring of the behaviour and demonstration of the behaviour).

Eight opportunities for intervention functions, utilised by the training to target key TDF domains, were missed and ten were seized. Intervention functions persuasion, restriction, and environmental restructuring would address more than one of the key TDF domains, and intervention functions incentivisation and coercion would address one. Supplementary Material 4 shows the percentage of relevant BCTs utilised by the MECC TtT programme. MECC TtT training only adequately addressed (> 50% of relevant BCTs utilised [37]) the key TDF domain skills (66.6% of relevant BCTs were utilised). At least one relevant BCT was utilised for domains intentions (33.3%), social influences (20%), knowledge (20%) and environmental context and resources (14.29%) and no relevant BCTs were utilised to address beliefs about consequences. Thus, recommendations for future MECC TtT training focused on implementing BCTs relevant to the five insufficiently addressed key TDF domains (see Table 9 for recommendations including amendments made after the second professional contributor meeting). BCTs that would be highly congruent with key TDF domains and are not currently utilised by MECC TtT training are Social Support (Practical), Information about Health Consequences (the only BCT to address three key TDF domains), Information about Social and Environmental Consequences, and Incentive (Outcome). Application of these four BCTs would therefore show the highest efficiency in addressing the most relevant barriers and facilitators to cascading MECC training.

**Table 9:**
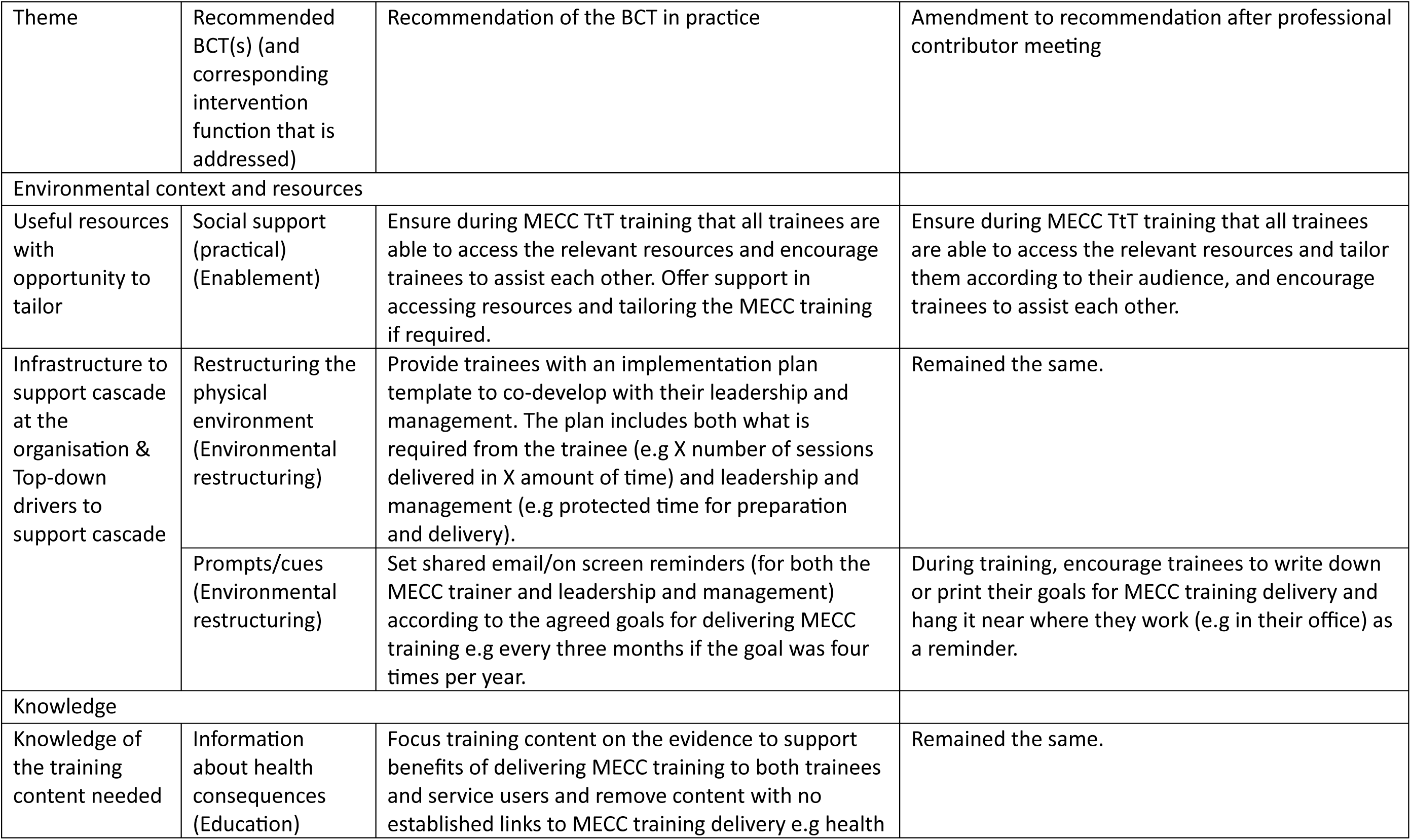
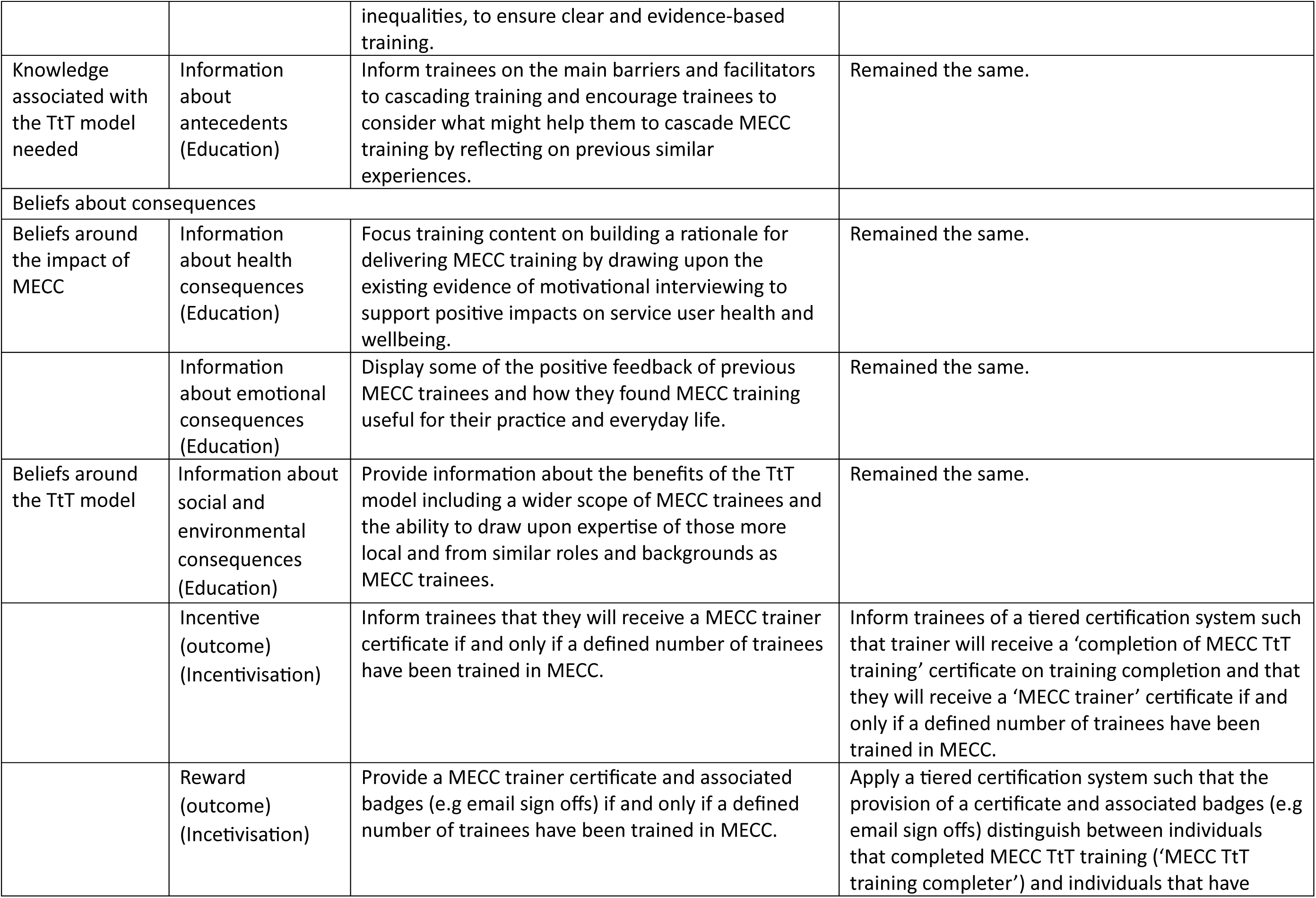
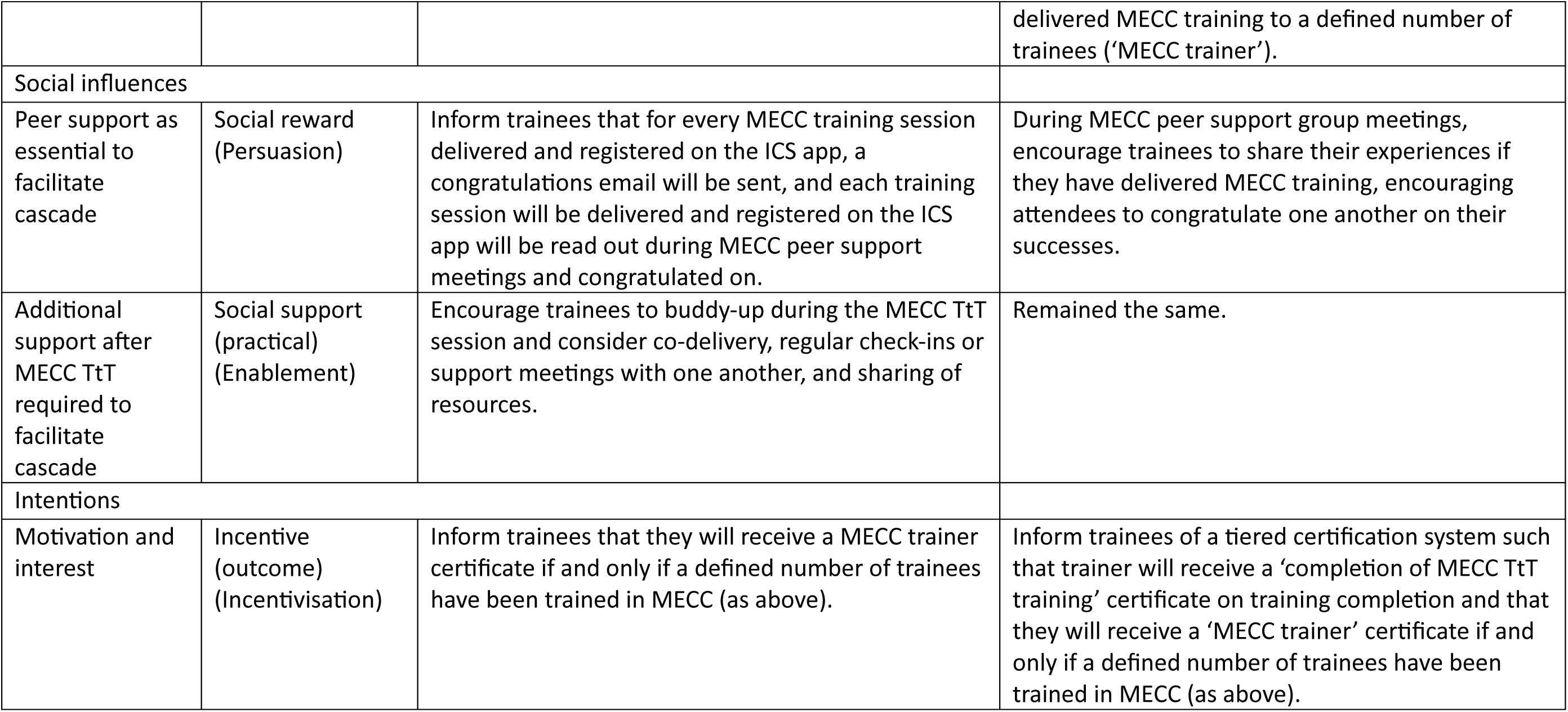
Recommended BCTs to adequately address key TDF domains that are not sufficiently addressed by the current MECC TtT training, informed by the qualitative interviews and quantitative data. Recommendations for each BCT in practice were assessed for appropriateness by the professional contributor panel and amended accordingly. Corresponding intervention functions are identified from Michie et al. (2014).

| Theme | Recommended BCT(s) (and corresponding intervention function that is addressed) | Recommendation of the BCT in practice | Amendment to recommendation after professional contributor meeting |
| --- | --- | --- | --- |
| Environmental context and resources |  |  |  |
| Useful resources with opportunity to tailor | Social support (practical) (Enablement) | Ensure during MECC TtT training that all trainees are able to access the relevant resources and encourage trainees to assist each other. Offer support in accessing resources and tailoring the MECC training if required. | Ensure during MECC TtT training that all trainees are able to access the relevant resources and tailor them according to their audience, and encourage trainees to assist each other. |
| Infrastructure to support cascade at the organisation & Top-down drivers to support cascade | Restructuring the physical environment (Environmental restructuring) | Provide trainees with an implementation plan template to co-develop with their leadership and management. The plan includes both what is required from the trainee (e.g X number of sessions delivered in X amount of time) and leadership and management (e.g protected time for preparation and delivery). | Remained the same. |
|  | Prompts/cues (Environmental restructuring) | Set shared email/on screen reminders (for both the MECC trainer and leadership and management) according to the agreed goals for delivering MECC training e.g every three months if the goal was four times per year. | During training, encourage trainees to write down or print their goals for MECC training delivery and hang it near where they work (e.g in their office) as a reminder. |
| Knowledge |  |  |  |
| Knowledge of the training content needed | Information about health consequences (Education) | Focus training content on the evidence to support benefits of delivering MECC training to both trainees and service users and remove content with no established links to MECC training delivery e.g health inequalities, to ensure clear and evidence-based training. | Remained the same. |
| Knowledge associated with the TtT model needed | Information about antecedents (Education) | Inform trainees on the main barriers and facilitators to cascading training and encourage trainees to consider what might help them to cascade MECC training by reflecting on previous similar experiences. | Remained the same. |
| Beliefs about consequences |  |  |  |
| Beliefs around the impact of MECC | Information about health consequences (Education) | Focus training content on building a rationale for delivering MECC training by drawing upon the existing evidence of motivational interviewing to support positive impacts on service user health and wellbeing. | Remained the same. |
|  | Information about emotional consequences (Education) | Display some of the positive feedback of previous MECC trainees and how they found MECC training useful for their practice and everyday life. | Remained the same. |
| Beliefs around the TtT model | Information about social and environmental consequences (Education) | Provide information about the benefits of the TtT model including a wider scope of MECC trainees and the ability to draw upon expertise of those more local and from similar roles and backgrounds as MECC trainees. | Remained the same. |
|  | Incentive (outcome) (Incentivisation) | Inform trainees that they will receive a MECC trainer certificate if and only if a defined number of trainees have been trained in MECC. | Inform trainees of a tiered certification system such that trainer will receive a 'completion of MECC TtT training' certificate on training completion and that they will receive a 'MECC trainer' certificate if and only if a defined number of trainees have been trained in MECC. |
|  | Reward (outcome) (Incentivisation) | Provide a MECC trainer certificate and associated badges (e.g email sign offs) if and only if a defined number of trainees have been trained in MECC. | Apply a tiered certification system such that the provision of a certificate and associated badges (e.g email sign offs) distinguish between individuals that completed MECC TtT training ('MECC TtT training completer') and individuals that have delivered MECC training to a defined number of trainees ('MECC trainer'). |
| Social influences |  |  |  |
| Peer support as essential to facilitate cascade | Social reward (Persuasion) | Inform trainees that for every MECC training session delivered and registered on the ICS app, a congratulations email will be sent, and each training session will be delivered and registered on the ICS app will be read out during MECC peer support meetings and congratulated on. | During MECC peer support group meetings, encourage trainees to share their experiences if they have delivered MECC training, encouraging attendees to congratulate one another on their successes. |
| Additional support after MECC TtT required to facilitate cascade | Social support (practical) (Enablement) | Encourage trainees to buddy-up during the MECC TtT session and consider co-delivery, regular check-ins or support meetings with one another, and sharing of resources. | Remained the same. |
| Intentions |  |  |  |
| Motivation and interest | Incentive (outcome) (Incentivisation) | Inform trainees that they will receive a MECC trainer certificate if and only if a defined number of trainees have been trained in MECC (as above). | Inform trainees of a tiered certification system such that trainer will receive a 'completion of MECC TtT training' certificate on training completion and that they will receive a 'MECC trainer' certificate if and only if a defined number of trainees have been trained in MECC (as above). |

### Triangulation of data

Only motivation and confidence significantly predicted Intentions, and even then, the model only accounted for a small amount (16.7%) of the variance in intentions.

Findings from the SBA indicated that change in Skills and Beliefs about Consequences may also predict Intentions, explaining some of the variance. Furthermore, many of the influential factors in determining cascade of MECC training concern after the MECC TtT training; Environmental Context and Resources is likely the most influential domain, followed by Social Influences. Although knowledge of the content significantly increased it did not significantly predict intentions. Knowledge refers to knowledge around MECC whereas the qualitative findings show that the current training is less proficient in informing attendees on the TtT model for MECC training and how it operates.

It was clear from the post survey data, interviews, and post survey comments that individuals were highly positive about the MECC TtT programme and generally reported high satisfaction with their training experience. Particularly, attendees showed positive affect towards the principal trainer noting characteristics such as *‘approachable’*, *‘knowledgeable’*, *‘inclusive’*, and an ability to make attendees feel *‘at ease’*. Negative feedback instead centred around a lack of guidance on *‘what next’* and overwhelm at the course content. Perhaps relatedly, the quantitative data indicate that intentions most often do not translate into cascade behaviour, suggesting that participants initially feel positive and motivated but soon encounter challenges that prevents cascade combined with a lack of guidance and support to overcome them. Also, the post survey comments indicated that many participants clearly left MECC with a new or renewed sense that MECC was valuable, no significant changes in motivation scores indicated participants were less motivated to deliver or were unaware of their role in delivering MECC training.

Some participants with an overseeing role of MECC implementation within their organisation reported that rate of cascade was so low that they had reduced or stopped rollout of TtT completely. Indeed, the quantitative data on fidelity supports the failure of the TtT model of MECC to deliver MECC training at scale. A theme that spanned across TDF domains was that training delivery was perceived to become easier with repetition; that confidence would grow, that content would become easier to recall, and that delivery would improve. However, as the quantitative data indicates, most trainees never reach the point of repetition.

## Discussion

This study aimed to explore the nature of MECC training cascade within the NENC and how to best utilise facilitators and overcome barriers. The fidelity data confirmed training cascade as problematic; those employed to deliver training (the principal trainer coordinated by the Regional Delivering MECC at Scale coordinator) continued to act as the primary providers of MECC training. According to the available data there were only seven MECC TtT trainers that had delivered more than two sessions. Furthermore, some participants reported reducing or discontinuing implementation of the TtT model within their organisation due to lack of success. Triangulation of the available data suggested that current training was insufficient in providing TtT trainees with the skills and thus confidence to deliver training, and that further support was required to determine success, particularly from leadership and management which also largely determined motivation to cascade MECC training. Also, TtT trainees varied greatly in their existing knowledge and skills, incongruent with the current homogenous approach to MECC TtT training. Thus, to deliver an efficient model of MECC training delivery as intended, access to MECC training alongside MECC TtT training, more selectiveness in who attends MECC TtT training, and tailored levels of support during and post training should be ensured. Furthermore, the current MECC TtT training can be optimised by including practice of training delivery, problem solving, collaboratively setting goals for cascade, and planning delivery of MECC training, such as creating an implementation plan.

When compared to the previous quantitative study of a MECC TtT programme comparing pre and post evaluation surveys [27], MECC TtT based on the HCS framework successfully increased numerous of the key identified domains within the current study (Knowledge, Skills, Intentions, Environmental Context and Resources, Social Influences) so that three of the domains were no longer barriers to cascade (Knowledge, Beliefs about Consequences and Intentions). In contrast to the NENC approach, HCS TtT training is more intensive (one full day), is delivered additionally and separately to the HCS training, is non-didactic, provides a training manual with an explanation of the purpose of and detailed guidance on conducting each activity, and involves practice of training delivery [27]; all of which appear to facilitate cascade based on findings from the current study. Thus, HCS TtT is recommended as an existing training package that would address some of the main barriers to cascade of MECC training. However, the findings from Hollis et al. [27] also indicate that some key TDF domains may be best targeted after the training and within an individual’s organisation (Environmental Context and Resources and Social Influences) and that it may not be feasible for the adequate skills required for training delivery to be translated within one day, supporting the findings of the current study that previous training experience was advantageous. The MECC literature regarding the TtT model has also identified the importance of top-down support [18, 28], creating designated roles for implementation [18], and making expectations of cascade clear at the sign-up stage of training [18]. Particularly, the current study found full and involved support form leadership and management to be one on the most fundamental facilitators to cascade of MECC training. Specific strategies common with those suggested within the current study include a buddy-up system of training delivery [18, 27], refresher sessions [27], support network meetings [18, 27], and provision of incentives and rewards for cascading MECC training to encourage trainer motivation [28]. However, instead of a need for additional resources [27], the current study identified a need for support in navigating existing resources.

The percentage of trained trainers that reported cascade of MECC training was extremely low (4.4%), compared to more successful TtT programmes [21, 23, 55] with most trainers delivering training repeatedly [23, 55]. Comparing the current TtT model to more successful examples of implementation of TtT training [20, 21, 23, 55], successful TtT models possessed a more intensive focus on development of training and facilitation skills [21, 23, 55], centered around problem solving, interaction, and reflection [23], focused on how to plan and organise training [21], required demonstration of competency to deliver training [55], separated the original training and the TtT component [23, 55], provided funding to plan and prepare to deliver training [23], included practice of training delivery [20, 21, 23, 55], and included preparation before the workshop [21]. Furthermore, after training sessions, successful TtT models provided a nominated contact for technical assistance after TtT training if practical planning was not included within the training sessions [20, 23], only provided a certificate of being a certified trainer on successful cascade of training [23], built in peer support including a ‘debrief’ after delivery of training [23], provided support for tailoring whilst retaining quality and consistency of the training content [20], and provided follow-up or ‘refresher’ sessions [20]. Similarly to the current study, individuals that successfully cascaded training were most likely to be from roles that indicated that they were a lead or manager of the training topic or were professional trainers [23]. Additional suggested strategies by the current study and common with the wider TtT literature include scope for training to be tailored with advice on how to do so [46, 56], co-production of resources for trainers [46], rewards for training delivery [48], buddy-up systems to deliver training [24], designated time for staff to deliver and attend training [48], and ongoing support from the principal trainer [21, 46].

One key factor in the current study was that the TtT training was accessible to anyone aware of the training, whereas successful examples of implementation of the TtT model are far more selective. For example, one study described that 265 individuals completed an application for the TtT program and only 103 were judged to be eligible [23]. Subsequently, 59% of the 87 individuals that completed the training successfully cascaded training, most of whom indicated that they would continue to deliver training [23]. Previous examples of trainer selection included nominated individuals [20, 55] such as individuals that demonstrated potential as trainers [20], or current trainers that possessed previous training experience, relevant knowledge of the topic, and a willingness to deliver training [23, 55], as well as a letter of support from the individual’s organisation [23]. As suggested within the current study, evidence of involved top-down buy-in from leadership and management may also be demonstrated through the completion of a co-produced implementation plan for MECC training (see Table 9). Indeed, within successful examples of the TtT model attendees felt significantly more prepared and skillful at presenting and public speaking after training when compared to before and most felt that they understood how to conduct training [23]. Thus, refining the accessibility of MECC TtT training may dramatically increase its efficiency. Furthermore, the TDF domain Environmental Context and Resources was identified in the current study to be by far the most relevant domain in determining successful cascade of MECC training, although the ability to address this domain is inevitably limited by the available resources of implementers of the TtT model. Thus, the lower the resources provided by the training programme, the more selective the inclusion criteria for the TtT training must be to ensure individuals are sufficiently equipped to deliver training. However, as demonstrated in the successful example by Lang et al. [23], ideally a TtT model would be high in both. When both of these key barriers are addressed, the main barrier becomes recruiting attendees of training [23], and the current study indicated no lack of interest in MECC training.

### Strengths and limitations

Given that all quantitative data was secondary, the authors had no influence over what and how data was collected, and thus the data was not comprehensive meaning the potential for bias was high. The estimated rate of MECC training cascade was dependent on data as entered into the ICS application, and indeed some interview participants noted a preference to recording cascade data outside of the application for their own organisation’s benefit instead. Furthermore, any training cascade outside of the regional delivery was likely not captured by the current evaluation. Whilst the authors named variables of Motivation, Confidence, and Knowledge, a single item informed each variable and none were theoretically based. The small proportion of variance in Intentions accounted for indicates that the variables were not comprehensive or that there were other factors that influenced intention to cascade that were not measured by the current evaluation survey. A robust survey design could be guided by the TDF, as utilised within the previously mentioned MECC TtT survey [27]. Also, pre and post survey questions sometimes lacked clarity over whether they referred to delivering MECC or MECC training, and there was a lack of guidance on how participants should respond, further limiting conclusions that could be drawn from the quantitative data. Finally, variables of Confidence, Motivation, and Knowledge all demonstrated a ceiling effect such that any changes were more difficult to detect. Whilst there was a low response rate to post surveys, this was to be expected given that the active implementation of MECC TtT meant that recent attendees were yet to complete it.

One reflection on the content analysis of training was that coding of BCTs and intervention functions only identified whether they were present or not, rather than their degree of application. For example, although attendees were instructed to practice training delivery, there was no opportunity within the training session to do so. Also, although attendees were instructed to deliver a specific number of MECC training sessions within a specific timeframe, the quality of goal setting could have been improved by creating goals collaboratively with attendees so that goals were bespoke and perceived as realistic and achievable to the individual. To address this limitation, future research utilising the SBA method may benefit from also coding the degree of presence of BCTs and intervention functions and incorporating this measure into the subsequent mapping against key TDF domains.

### Conclusion

The implementation of the MECC TtT programme in the NENC was highly inefficient and did not facilitate the goal of delivering MECC at scale. Attendance at the MECC TtT programme was insufficient to determine cascade, and attendees depended on existing knowledge and skills and required additional support from programme providers and staff at their respective organisations to cascade MECC training. The current study identified strategies to implement both within and outside of MECC TtT training to improve future cascade.

Inclusion criteria for attendees of MECC TtT training should include those with existing training experience and knowledge of MECC, time to deliver MECC training, existing intentions and a plan of how and when to deliver MECC training, motivation to deliver training, awareness of expectations of cascade, and those for whom MECC training is part of their role or at least those with proactive support from their leadership and management.

TtT training should be separate and additional to core MECC training, build a rationale for delivering MECC training, and provide more comprehensive skills training including opportunity to practice training delivery. Meanwhile, to strengthen the ability to provide a rationale for the delivery of MECC training, researchers should focus on building the evidence base and subsequent rationale for MECC, including monitoring service user outcomes of MECC delivery. Where feasible, the level of support provided during and after training should be tailored to the existing knowledge, confidence, and skills of the individual. Such support should include facilitation of peer support, technical support, continued check-ins for accountability, and refresher sessions for refreshed knowledge and motivation. Peer support is key for successful cascade and is organically facilitated by face-to-face training.

Furthermore, individuals for whom delivery of MECC training is not part of their role would benefit from the receipt of rewards and incentives for MECC training delivery. The lower the support that can be provided, the tighter the inclusion criteria for TtT attendees should be. In summary, the TtT model for MECC is recommended as appropriate provided the specificity of attendees and the level of support provided to selected attendees is increased.

## Data Availability

All data produced are available online at https://reshare.ukdataservice.ac.uk/857461/

## References

1. Nichol, B., Kemp, E., Wilson, R., Rodrigues, A. M., Hesselgreaves, H., Robson, C., Haighton, C Establishing an updated consensus on the conceptual and operational definitions of Making Every Contact Count (MECC) across experts within research and practice internationally: a Delphi Study. . Public Health, 2024.

2. Public Health England, P., Making Every Contact Count (MECC): Consensus Statement. 2016.

3. Public Health England, P., Making Every Contact Count (MECC): implementation guide. 2018.

4. Adam, L.M., et al., Use of healthy conversation skills to promote healthy diets, physical activity and gestational weight gain: Results from a pilot randomised controlled trial. Patient education and counseling, 2020. 103(6): p. 1134–1142.

5. DiClemente, C.C., et al., Motivational interviewing, enhancement, and brief interventions over the last decade: A review of reviews of efficacy and effectiveness. Psychology of Addictive Behaviors, 2017. 31(8): p. 862.

6. Nichol, B., A.M. Rodrigues, R. Wilson, and C. Haighton, A Systematic Review of the Effectiveness of Brief Health Behaviour Change Interventions on Service Users Accessing the Third and Social Economy Sector. Health & Social Care in the Community, 2023. 2023.

7. Rollnick, S. and W.R. Miller, What is motivational interviewing? Behavioural and cognitive Psychotherapy, 1995. 23(4): p. 325–334.

8. Watson, D., et al., Adapting Making Every Contact Count/Healthy Conversation skills to pilot online Supportive Conversations training in response to Covid-19. Behavioural Science & Public Health, 2020.

9. Black, C., et al., Healthy conversation skills: increasing competence and confidence in front-line staff. Public health nutrition, 2014. 17(3): p. 700–707.

10. Chisholm, A., et al., Online behaviour change technique training to support healthcare staff ‘Make Every Contact Count’. BMC health services research, 2020. 20(1): p. 1–11.

11. Lawrence, W., et al., Meeting the UK Government’s prevention agenda: primary care practitioners can be trained in skills to prevent disease and support self-management. Perspectives in Public Health, 2020: p. 1757913920977030.

12. Jarman, M., et al., Healthy conversation skills as an intervention to support healthy gestational weight gain: Experience and perceptions from intervention deliverers and participants. Patient education and counseling, 2019. 102(5): p. 924–931.

13. Lawrence, W., et al., ‘Making every contact count’: evaluation of the impact of an intervention to train health and social care practitioners in skills to support health behaviour change. Journal of health psychology, 2016. 21(2): p. 138–151.

14. Lawrence, W., et al., How can we best use opportunities provided by routine maternity care to engage women in improving their diets and health? Maternal & child nutrition, 2020. 16(1): p. e12900.

15. Hollis, J.L., et al., The impact of Healthy Conversation Skills training on health professionals’ barriers to having behaviour change conversations: a pre-post survey using the Theoretical Domains Framework. BMC health services research, 2021. 21(1): p. 1–13.

16. Parchment, A., et al., How useful is the Making Every Contact Count Healthy Conversation Skills approach for supporting people with musculoskeletal conditions? Journal of Public Health, 2022: p. 1–17.

17. Parchment, A., et al., ‘I can feel myself coming out of the rut’: a brief intervention for supporting behaviour change is acceptable to patients with chronic musculoskeletal conditions. BMC Musculoskeletal Disorders, 2023. 24(1): p. 1–12.

18. Rodrigues, A.M., et al., Mapping regional implementation of ‘Making Every Contact Count’: mixed methods evaluation of implementation stage, strategies, barriers and facilitators of implementation. BMJ Open, 2024. 14(7): p. e084208.

19. Servey, J., et al., The ripple effect: A train-the-trainer model to exponentially increase organizational faculty development. MedEdPublish, 2020. 9.

20. Yarber, L., et al., Evaluating a train-the-trainer approach for improving capacity for evidence-based decision making in public health. BMC health services research, 2015. 15: p. 1–10.

21. Tobias, C.R., A. Downes, S. Eddens, and J. Ruiz, Building blocks for peer success: lessons learned from a train-the-trainer program. AIDS patient care and STDs, 2012. 26(1): p. 53–59.

22. Orfaly, R.A., et al., Train-the-trainer as an educational model in public health preparedness. Journal of Public Health Management and Practice, 2005. 11(6): p. S123–S127.

23. Lang, J., et al., Building capacity for workplace health promotion: Findings from the Work@ Health® Train-the-Trainer program. Health promotion practice, 2017. 18(6): p. 902–911.

24. Lloyd, B., L. Rychetnik, M. Maxwell, and T. Nove, Building capacity for evidence-based practice in the health promotion workforce: evaluation of a train-the-trainer initiative in NSW. Health Promotion Journal of Australia, 2009. 20(2): p. 151–154.

25. Burr, C.K., D.S. Storm, and E. Gross, A faculty trainer model: increasing knowledge and changing practice to improve perinatal HIV prevention and care. AIDS Patient Care & STDs, 2006. 20(3): p. 183–192.

26. Trabeau, M., et al., A comparison of “Train-the-Trainer” and expert training modalities for hearing protection use in construction. American journal of industrial medicine, 2008. 51(2): p. 130–137.

27. Hollis, J.L., et al., Evaluating a train-the-trainer model for scaling-up healthy conversation skills training: a pre-post survey using the theoretical domains framework. Patient Education and Counseling, 2022. 105(10): p. 3078–3085.

28. Turner, R., et al., Experiences of implementing the ‘Making Every Contact Count’initiative into a UK integrated care system: an interview study. Journal of Public Health, 2023: p. fdad173.

29. Corelli, R.L., et al., Evaluation of a train-the-trainer program for tobacco cessation. American journal of pharmaceutical education, 2007. 71(6).

30. Farquhar, M.C., G. Ewing, and S. Booth, Using mixed methods to develop and evaluate complex interventions in palliative care research. Palliative medicine, 2011. 25(8): p. 748–757.

31. Craig, P., et al., Developing and evaluating complex interventions: the new Medical Research Council guidance. Bmj, 2008. 337.

32. Dusenbury, L., R. Brannigan, M. Falco, and W.B. Hansen, A review of research on fidelity of implementation: implications for drug abuse prevention in school settings. Health education research, 2003. 18(2): p. 237–256.

33. Bull, E.R. and H. Dale, Improving community health and social care practitioners’ confidence, perceived competence and intention to use behaviour change techniques in health behaviour change conversations. Health & Social Care in the Community, 2021. 29(1): p. 270–283.

34. Michie, S., L. Atkins, and R. West, The behaviour change wheel: A guide to designing interventions. 1st ed. 2014, Great Britain: Silverback Publishing. 1003-1010.

35. Cane, J., D. O’Connor, and S. Michie, Validation of the theoretical domains framework for use in behaviour change and implementation research. Implementation science, 2012. 7(1): p. 1–17.

36. Michie, S., et al., The behavior change technique taxonomy (v1) of 93 hierarchically clustered techniques: building an international consensus for the reporting of behavior change interventions. Annals of behavioral medicine, 2013. 46(1): p. 81–95.

37. Haighton, C., et al., Optimising making every contact count (MECC) interventions: a strategic behavioural analysis. Health Psychology, 2021.

38. Driscoll, D.L., A. Appiah-Yeboah, P. Salib, and D.J. Rupert, Merging qualitative and quantitative data in mixed methods research: How to and why not. 2007.

39. Nichol, B., Rodrigues, A. M., Audsley, S., Haste, A., Tang, M. Y., Robson, C., Harland, J., Haighton, C., Exploring the ‘train the trainer’ model for delivering Making Every Contact Count (MECC) training at scale: A qualitative study. (under review).

40. Levitt, H.M., et al., Journal article reporting standards for qualitative primary, qualitative meta-analytic, and mixed methods research in psychology: The APA Publications and Communications Board task force report. American Psychologist, 2018. 73(1): p. 26.

41. Kotsis, S.V. and K.C. Chung, Application of see one, do one, teach one concept in surgical training. Plastic and reconstructive surgery, 2013. 131(5): p. 1194.

42. Faul, F., E. Erdfelder, A. Buchner, and A.-G. Lang, Statistical power analyses using G* Power 3.1: Tests for correlation and regression analyses. Behavior research methods, 2009. 41(4): p. 1149–1160.

43. Corp, I., IBM SPSS Statistics for Windows, Version 28.0. 2021, IBM Corp: Armonk, NY.

44. Malterud, K., V.D. Siersma, and A.D. Guassora, Sample size in qualitative interview studies: guided by information power. Qualitative health research, 2016. 26(13): p. 1753–1760.

45. Michie, S., et al., Making psychological theory useful for implementing evidence based practice: a consensus approach. BMJ Quality & Safety, 2005. 14(1): p. 26–33.

46. Hayes, D., Cascade training and teachers’ professional development. ELT journal, 2000. 54(2): p. 135–145.

47. Wessex, H.E.E., Making Every Contact Count Toolkit. n.d.

48. Levine, S.A., et al., Practicing physician education in geriatrics: Lessons learned from a train-the-trainer model. Journal of the American Geriatrics Society, 2007. 55(8): p. 1281–1286.

49. Öztek, Z. and K. Angela, A new training approach for vaccinators: Cascade plus training. Turkish Journal of Public Health, 2022. 20(1): p. 164–176.

50. Mclaughlin, M., et al., Evaluating digital program support for the physical activity 4 everyone (PA4E1) school program: Mixed methods study. JMIR pediatrics and parenting, 2021. 4(3): p. e26690.

51. Ltd, Q.I.P., Nvivo. 2018.

52. Nichol, B., Rodrigues, A. M., Haighton, C., Transcripts of those who attended and had not attended the Making Every Contact Count (MECC) Train the Trainer programme in the North East and North Cumbria., U.D. Service, Editor. 2024: Colchester, Essex.

53. Tashakkori, A., C. Teddlie, and C.B. Teddlie, Mixed methodology: Combining qualitative and quantitative approaches. Vol. 46. 1998: sage.

54. Cohen, J., Statistical power analysis for the behavioral sciences (2nd ed.). 1988, New York: Psychology Press.

55. Hiner, C.A., et al., Effectiveness of a training-of-trainers model in a HIV counseling and testing program in the Caribbean Region. Human Resources for Health, 2009. 7: p. 1–8.

56. Kienlin, S., et al., Ready for SDM: evaluating a train-the-trainer program to facilitate implementation of SDM training in Norway. BMC Medical Informatics and Decision Making, 2021. 21(1): p. 1–19.

