## Supplementary Material for "Mixed methods evaluation of the “train the trainer” model for delivering core “Making Every Contact Count” (MECC) training"

**Supplementary Material 1:** Pre and post survey evaluation questions included in the analysis. Type of question/scale is indicated in brackets. All questions are presented in the same order that they were presented to attendees. Italics marks the variable names included in the analysis.

Pre survey

*Motivation:* How important is it for you to deliver MECC training? (Likert, 1-5)

*Knowledge:* How would you rate your level of awareness of MECC? (Likert, 1-5)

How do you feel before this training session? (Grid of words; Confident, Uncertain, Energised, Competent, Apprehensive, Enthused, Knowledgeable, Confused, Frustrated, Motivated, Positive, Focused)

*Confidence:* How confident do you feel in having a training session about MECC? (Likert, 1-5)

Post survey

*Motivation:* How important is it for you to deliver MECC training? (Likert, 1-5)

*Confidence:* How confident do you feel in delivering MECC training? (Likert, 1-5)

How do you feel after this training session? (Grid of words; Confident, Uncertain, Energised, Competent, Apprehensive, Enthused, Knowledgeable, Confused, Frustrated, Motivated, Positive, Focused)

Is there anything you would change within the training or any further information that would help? (free text)

*Knowledge:* How would you rate your level of awareness of MECC? (Likert, 1-5)

What did you find most useful from today’s training? (free text)

*Readiness:* How ready are you to deliver training? (Likert, 1-5)

What will you change in your practice as a result of this training? (free text)

*Intention:* How many courses do you plan to deliver in the next 12 months? (free text)

Is there anything else you would like to tell us about today’s training? (free text)

If you require some support, what support do you require? (free text)

*Trainer: positive response:* To what extent did your trainer respond positively to being asked questions? (Likert, 1-5)

*Trainer: enthusiastic:* To what extent did your trainer speak with an enthusiastic tone? (Likert, 1-5)

*Trainer: positive language:* To what extent did your trainer use language to praise, support and show positive regard to learners? (Likert, 1-5)

*Trainer: listened:* To what extent did your trainer listen to learners? (Likert, 1-5)

*Trainer: eye contact:* To what extent did your trainer make eye contact with multiple learners? (Likert, 1-5)

*Trainer: used names:* To what extent did your trainer use the names of the learners? (Likert, 1-5)

*Trainer: curiosity in topic:* To what extent did your trainer demonstrate their own curiosity and interest in the topic? (Likert, 1-5)

*Trainer: learning into practice:* To what extent did your trainer help the learners think about how they might put their learning into practice when they go back to work? (Likert, 1-5)

What is the most important thing you got from your MECC training? (free text)

Any comments or suggestions that may help us to improve the training? (free text)

**Supplementary Material 2:** Assumptions testing for selecting the appropriate statistical tests for the quantitative analyses.

Differences pre and post survey:

On testing normality assumptions for differences pre and post training, all differences were highly significant when tested using the Shapiro Wilk test (p < .001). Motivation and Confidence showed an s-shaped curve on the resulting Q-Q plots and Knowledge showed too high a proportion of outliers (n = 22, 5.9%). Thus, a non-parametric test was selected as most appropriate.

Comparison between baseline of completers and non-completers:

The data was ordinal and not normally distributed, showing statistical significance for all groups at the *p* < .001 level when the Shapiro Wilk test was applied. Homogeneity of variance was significantly different for Confidence (Levene’s statistic = 8.53, *p* = .004) but not Knowledge (Levene’s statistic = 1.37, *p* = .243) or Motivation (Levene’s statistic = 3.50, *p* = .062) when Levene’s test based on median was applied. A legacy procedure for each of the three dependent variables confirmed that the distribution followed the same shape for completers and non-completers, thus a comparison of medians was appropriate (non-parametric analyses).

Sample size calculation for ordinal regression:

Two post survey questions assessing Intention and Readiness provided the potential to enter as the dependent variable into a predictive model. Due to the small likert scale, the latter question required an ordinal regression. To calculate required sample size to provide sufficient statistical power to detect a significant effect, the general rule of thumb provided by Peduzzi et al. was applied (1), which states around 10 cases of each level of the dependent variable are required for each independent variable. Applying this rule, entering mode of delivery and post survey Knowledge, Confidence, and Motivation as predictors, 40 cases are required of scale points one to five. This was not satisfied for all three variables (Knowledge, Confidence, and Motivation)- scale points one and five did not fulfil this criteria.

Multiple linear regression:

Assumptions for a multiple linear regression were tested. There was independence of residuals (Durbin-Watson statistic = 2.063), the relationships between the independent variables and the dependent variables appeared to be linear, and there was homoscedasticity, as assessed by visual inspection of a plot of studentized deleted residuals versus unstandardized predicted values. Furthermore, none of the independent variables were highly correlated (> .7) with one another) and all tolerance values were above .1, providing no concerns with collinearity of independent variables. Ten outliers were identified based on standardized residuals that were greater than ±3 standard deviation, all of which were for participants who intended to conduct between 8-10 sessions, indicating a lack of fit of the model to predicting extreme values. However, no outliers based on standardized deleted residuals ±3 standard deviations were identified. Leverage values were also considered to be low risk (< .2) (2) and there were no Cook’s values higher than one (3). Inspection of the histogram of standardised residuals and P-P plot indicated the distribution was normal enough to proceed, although the normal distribution was skewed by outliers of higher values.

References

1. Peduzzi P, Concato J, Kemper E, Holford TR, Feinstein AR. A simulation study of the number of events per variable in logistic regression analysis. Journal of clinical epidemiology. 1996 Dec 1;49(12):1373-9.
2. Huber PJ. Robust statistics. New York: John Wiley & Sons; 1981.
3. Cook RD, & Weisberg, S. Residuals and influence in regression. New York: Chapman & Hall; 1982.

**Supplementary Material 3:** assumptions testing for the sensitivity analysis of the multiple linear regression showed there was independence of residuals (Durbin-Watson statistic = 1.056), scatter plots indicated a linear relationship of independent variables with the dependent variable, none of the independent variables were highly correlated (> .7) with one another) and all tolerance values were above .1, providing no concerns with collinearity of independent variables. Leverage values were also considered to be low risk (< .2) (ref Huber 1981) and there were no Cook’s values higher than one (ref Cook and Weisberg, 1982). Inspection of the histogram of standardised residuals and P-P plot indicated a normal distribution.

The table below shows regression coefficients and standardised errors for the multiple linear regression analysis to predict number of MECC training sessions intended to deliver, when removing 15 participants who intended to deliver ≥ 8 sessions. B = unstandardised regression coefficient, CI = confidence interval (LL = lower limit, UL = upper limit) at α = .05, SE B = standard error of the coefficient, β = standardised coefficient.

| Independent variable | *B* | 95% CI for B |  | SE *B* | β |
| --- | --- | --- | --- | --- | --- |
|  |  | LL | UL |  |  |
| Mode of delivery | .031 | -.330 | .391 | .183 | .008 |
| Knowledge | .038 | -.203 | .279 | .123 | .019 |
| Motivation | .431 | .259 | .602 | .087 | .290 |
| Confidence | .348 | .157 | .540 | .097 | .210 |

**Supplementary Material 4:** The utilisation of relevant BCTs to the six key TDF domains. All relevant BCTs for each domain (aside from Social/ professional role and identity, of which there are no linked BCTs) are listed alongside whether they are utilised by the MECC TtT training. Utilised BCTs are underlined.

| BCT paired with key TDF domains |  | Proportion of BCTs utilised (%) |
| --- | --- | --- |
| Environmental context and resources  Social support (practical)  Prompts/cues  Remove aversive stimulus  Restructuring the physical environment  Restructuring the social environment  Avoidance/reducing exposure to cues for the behaviour  Adding objects to the environment |  | 14.29 |
| Knowledge  Biofeedback  Instruction on how to perform behaviour  Information about antecedents  Information about health consequences  Information about social and environmental consequences |  | 20 |
| Skills  Instruction on how to perform behaviour  Behavioural practice/rehearsal  Graded tasks |  | 66.7 |
| Beliefs about consequences  Information about health consequences  Salience of consequences  Information about social and environmental consequences  Anticipated regret  Information about emotional consequences  Pros and cons  Comparative imagining of future outcomes  Material incentive (behaviour)  Incentive (outcome)  Reward (outcome) |  | 0 |
| Social influences  Social support (unspecified)  Social support (practical)  Social comparison  Information about others’ approval  Social reward |  | 20 |
| Intentions  Goal setting (behaviour)  Information about health consequences  Incentive (outcome) |  | 33.3 |
